# ci-fGBD: Cluster-Integrated Fast Generalized Bruhat Decomposition for Multimodal Data Clustering in Alzheimer’s Disease

**DOI:** 10.1101/2025.09.03.25334761

**Authors:** Lokendra S. Thakur, Gurpreet Bharj, Lokesh Sangabattula, Bushra Malik

**Affiliations:** Innovation Engine Lab (iEL), MDtRI, Hyderabad, INDIA; Department of Neurology, Massachusetts General Hospital, Harvard Medical School Massachusetts, USA; Institute for Immunity, Transplantation, and Infection, Stanford University, Stanford, California, USA; Department of Materials Science, Massachusetts Institute of Technology, Cambridge, Massachusetts, USA; Neuroscience and Human Biology Program, Department of Biological Sciences, University of Toronto, Toronto, Ontario, CANADA

## Abstract

Multimodal biomedical datasets, such as those from neurodegenerative disease cohorts, present significant challenges in stratifying heterogeneous patient populations due to missing values, high dimensionality, and modality-specific biases. Traditional clustering methods often require extensive preprocessing and fail to integrate heterogeneous data types effectively. We introduce ci-fGBD **(Cluster-Integrated Fast Generalized Bruhat Decomposition)**, a novel matrix factorization and clustering framework that natively operates on block-structured, multimodal datasets. ci-fGBD extends the classical Bruhat decomposition by jointly learning latent representations and patient clusters while automatically harmonizing contributions across diverse modalities, including neuroimaging, cognitive assessments, genomics, wearable sensors, and environmental exposures. Benchmarking against standard methods on real datasets demonstrates that ci-fGBD consistently identifies clinically meaningful subgroups, capturing subtle biological, cognitive, and demographic heterogeneity in Alzheimer’s disease cohorts with superior interpretability and robustness.

## 1 Introduction

Recent advancements in precision medicine increasingly highlight the importance of identifying homogeneous patient subgroups within highly heterogeneous diseases such as Alzheimer’s Disease (AD).^**1, 2**^ Traditional clustering approaches, including k-means,^**3**^ hierarchical clustering,^**4**^ and Gaussian Mixture Models,^**5**^ are sensitive to preprocessing choices and often fail to integrate high-dimensional, multimodal datasets effectively.^**6, 7**^ While matrix decomposition techniques such as Principal Component Analysis (PCA)^**8**^ or Non-negative Matrix Factorization (NMF)^**9**^ can capture latent structures, they neither produce clusters natively nor handle structured missingness inherent in real-world datasets.^**10, 11**^ As a result, critical biological and clinical patterns, including subtle subtypes or demographic-specific trajectories, may remain undetected.

To address these limitations, we propose ci-fGBD, an unsupervised clustering framework based on **Fast Generalized Bruhat Decomposition (fGBD)**.^**12**^ ci-fGBD extends fGBD by embedding clustering constraints directly into the decomposition process, enabling joint learning of patient latent profiles and cluster assignments. The framework also incorporates an adaptive modality weighting mechanism, which balances contributions from diverse data types and prevents overrepresentation by any single modality. Together, these features make ci-fGBD robust, scalable, and interpretable, allowing for discovery of biologically and clinically meaningful subtypes across heterogeneous datasets, including the ADNI cohort.

## 2 Background and Motivation

### 2.1 Bruhat Decomposition and fGBD

The Bruhat decomposition is a canonical factorization technique from algebraic geometry that expresses matrices through a combination of permutation matrices and block-triangular structures.^**12**^ Its fast variant, fGBD, adapts these principles for data-intensive applications, enabling efficient block-wise decomposition of large matrices while accommodating missing entries and heterogeneous blocks. This approach is particularly suitable for complex biomedical datasets, such as those from ADNI^**13**^ and UK Biobank,^**14**^ where features span imaging, cognitive, genetic, and clinical modalities.

### 2.2 Need for Integrated Clustering

Although fGBD provides low-rank latent representations of patients and features, traditional implementations require a separate clustering step to identify patient subgroups, often resulting in misalignment between latent structure and cluster assignments. ci-fGBD overcomes this limitation by integrating clustering directly into the factorization process. By jointly optimizing latent profiles and cluster memberships while accounting for modality-specific contributions and missingness, ci-fGBD produces interpretable patient subtypes that are statistically coherent, biologically plausible, and clinically actionable. This integration ensures that downstream analyses, including cluster-specific cognitive trajectories, biomarker associations, and demographic insights, are robust, reproducible, and readily interpretable, forming a strong foundation for precision medicine applications.

## 3 Data Description

Data used in the preparation of this article were obtained from the Alzheimer’s Disease Neuroimaging Initiative (ADNI) database (adni.loni.usc.edu).^**15**^ The ADNI was launched in 2003 as a public-private partnership, led by Principal Investigator Michael W. Weiner, MD. The original goal of ADNI was to test whether serial magnetic resonance imaging (MRI), positron emission tomography (PET), other biological markers, and clinical and neuropsychological assessment can be combined to measure the progression of mild cognitive impairment (MCI) and early Alzheimer’s disease (AD). The current goals include validating biomarkers for clinical trials, improving the generalizability of ADNI data by increasing diversity in the participant cohort, and to provide data concerning the diagnosis and progression of Alzheimer’s disease to the scientific community. For up-to-date information, see adni.loni.usc.edu.

## 4 Methodology: ci-fGBD Framework

The **Cluster-Integrated Fast Generalized Bruhat Decomposition (ci-fGBD)** framework is designed to identify hidden disease subtypes from complex biomedical datasets, such as clinical assessments, neuroimaging, and molecular biomarkers. These datasets often have missing entries, which can hinder conventional analysis methods. Conceptually, ci-fGBD treats the dataset as a large table, with patients as rows and features as columns, and estimates missing values while summarizing each patient’s information into a compact, meaningful representation. This representation acts like a “fingerprint” for each patient, capturing essential disease characteristics in a lower-dimensional latent space.

The framework approximates the data as a product of two matrices:

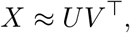

where *U* represents each patient in the latent space and *V* represents the contribution of each feature. Once patients are mapped into this space, they can be clustered to reveal disease subtypes, and the model adaptively weights different data modalities to avoid dominance by any single type of measurement.

### 4.1 Objective Function

The core of ci-fGBD is its joint factorization-clustering objective, formulated as Equation (1):

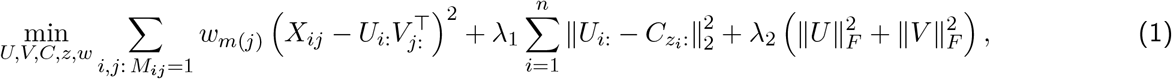

where *z*_*i*_ denotes the cluster assignment for patient *i, m*(*j*) identifies the modality of feature *j*, and *w* contains normalized weights for each modality. The parameters *λ*_1_ and *λ*_2_ control the influence of clustering and regularization.

In simple terms, this equation simultaneously achieves three important goals. The first term ensures that the model reconstructs the observed data as accurately as possible while handling missing values. The second term encourages patients assigned to the same cluster to have similar latent representations, which ensures that the clusters are meaningful and clinically relevant. The third term penalizes overly complex latent factors to prevent overfitting, which is particularly important in datasets with many features and relatively few patients. The weighting of each modality allows the framework to emphasize the most informative types of measurements, such as clinical tests or neuroimaging biomarkers. This balance is essential in Alzheimer’s disease research, where multimodal data integration is crucial to reveal latent disease subtypes that may correspond to different progression trajectories or therapeutic responses.

This formulation brings several novel advantages compared to traditional methods for clustering multimodal data in general and precisely in Alzheimer’s disease research. Unlike standard approaches that analyze each modality separately or rely on simple concatenation of features, **ci-fGBD jointly models all modalities**, capturing the interactions between clinical, imaging, and molecular data in a unified latent space. Its **pivot-free initialization** allows robust handling of missing values without introducing bias, which is critical for datasets with incomplete observations. The **adaptive weighting mechanism** ensures that no single modality dominates the analysis, giving each measurement type an appropriate influence on the resulting clusters. Furthermore, by combining latent factor representation with **cluster-aware learning**, ci-fGBD produces patient subtypes that are both statistically coherent and clinically interpretable, enabling the identification of meaningful disease subgroups and potential biomarkers for differential diagnosis or progression prediction. These innovations make the framework particularly suitable for complex diseases such as Alzheimer’s, Cancer, Diabetes studies, where heterogeneity and multimodal data integration are major challenges, and they distinguish ci-fGBD from previously published methods in the field.

### 4.2 Algorithmic Workflow

The ci-fGBD algorithm follows a structured workflow, summarized in Table. 1 and Figure. 1, to iteratively refine patient profiles and cluster assignments. The process begins with pivot-free initialization, which imputes missing values in a way that avoids bias from any single feature. Preliminary cluster assignments are then made based on these initial latent profiles using either k-means or pivot-free strategies. During the iterative optimization process, the latent patient profiles (*U*), feature contributions (*V*), cluster assignments (*z*), cluster centroids (*C*), and modality weights (*w*) are updated repeatedly. This refinement continues until convergence, meaning further updates do not significantly improve the objective function. Clinically, this iterative process is akin to gradually sharpening a blurred image: with each iteration, patient subtypes become more distinct and interpretable. The final outputs include patient latent profiles, cluster memberships, feature contributions, and a completed dataset with missing values imputed.

**Figure 1.**
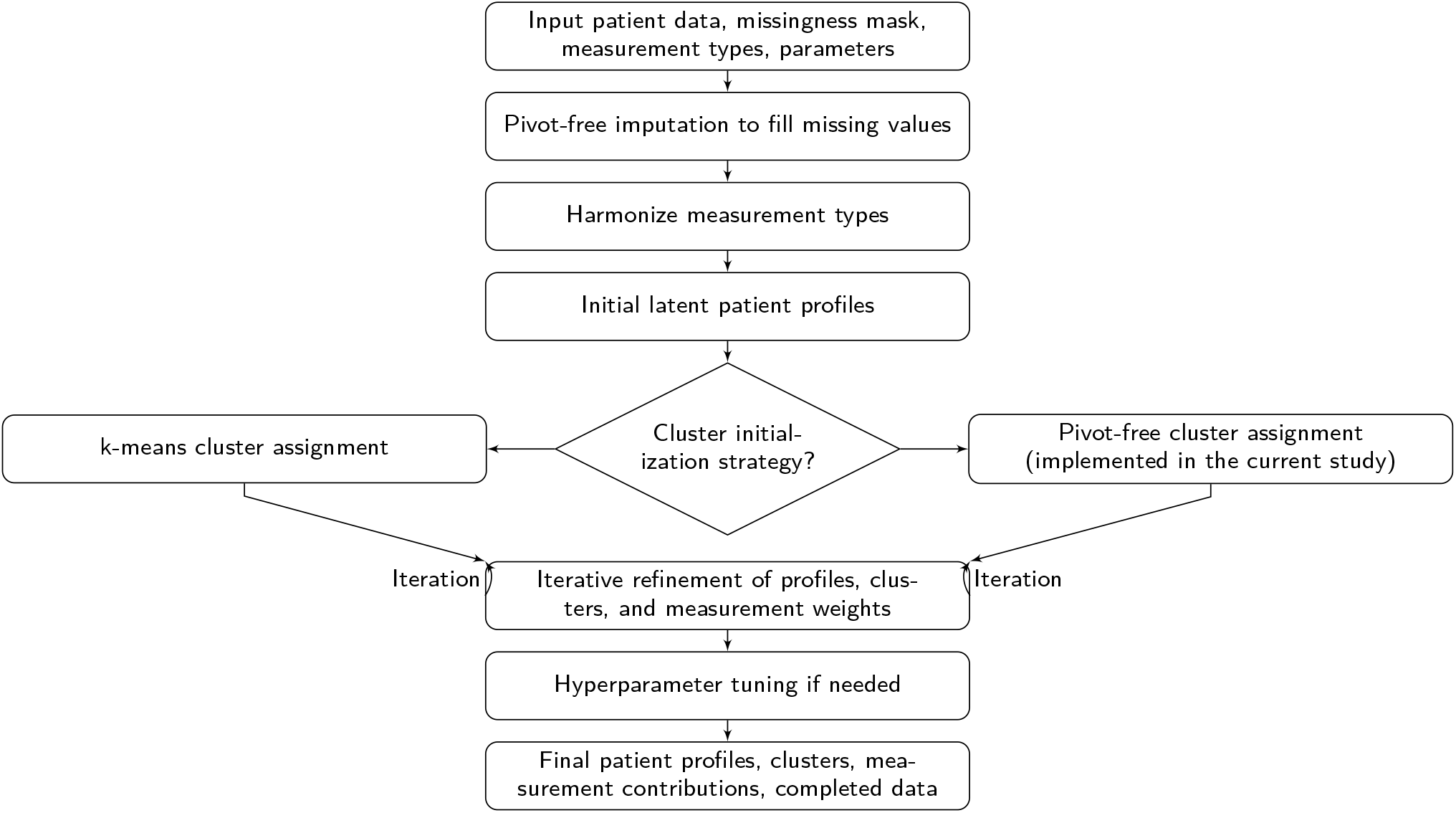
ci-fGBD pipeline showing the flow from raw patient data through imputation, latent profile calculation, cluster initialization, and iterative refinement to final outputs. Each stage is clinically interpretable, with iteration gradually revealing patient subtypes and key features.

### 4.3 Implementation Details

Table 1 maps each step of ci-fGBD to pseudocode and actual code, with explanations that highlight the clinical relevance of each stage. The pivot-free initialization provides unbiased latent representations and imputes missing data. Cluster initialization establishes starting groups for subtype identification, while iterative updates refine patient fingerprints, determine key features, and adjust modality weights. Convergence checks ensure that the process stops when further refinements are negligible. Optional hyperparameter tuning allows optimization of cluster numbers, latent dimensions, and regularization terms.

**Table 1.**
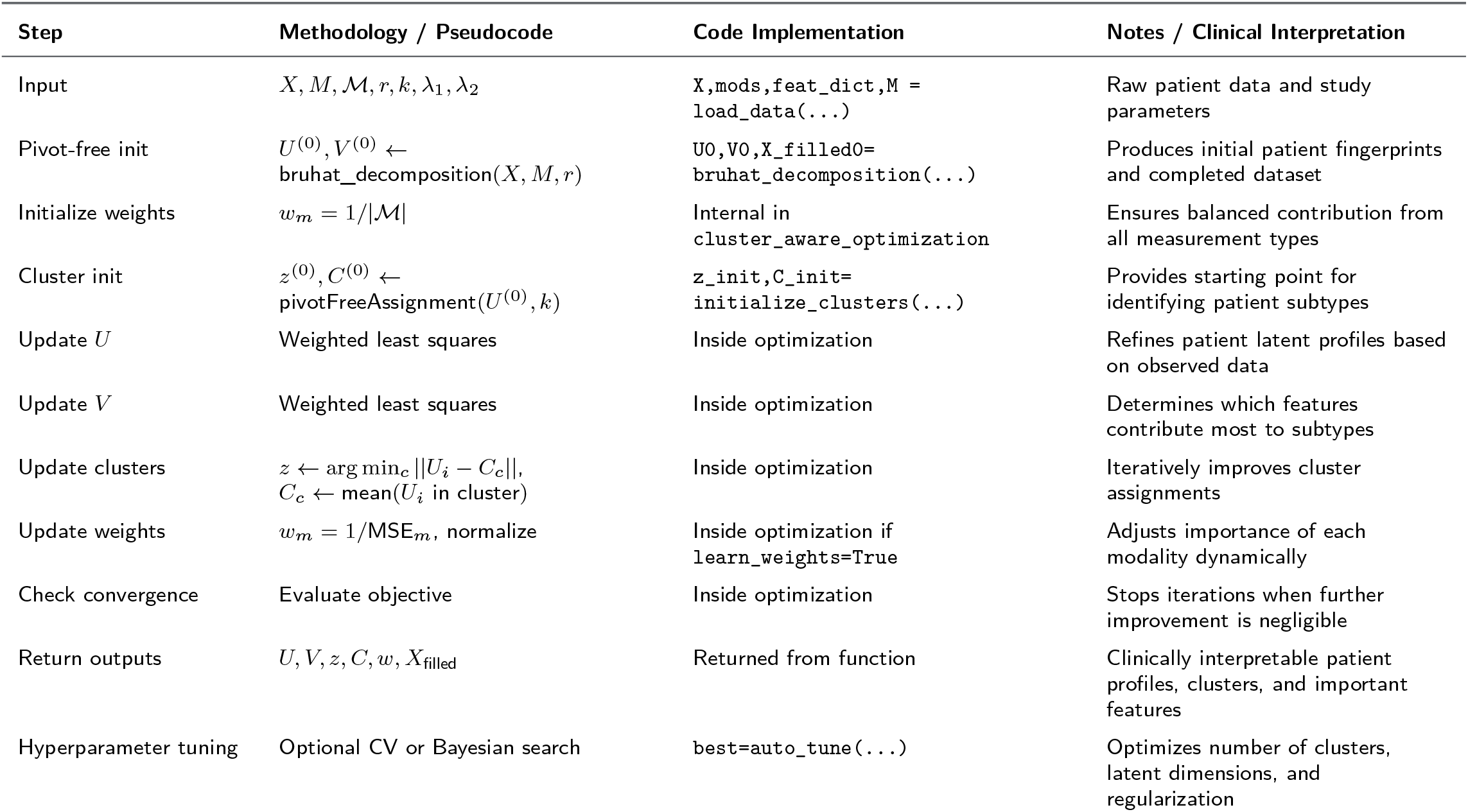
Mapping of ci-fGBD pseudocode to code implementation with clinical interpretation.

**Table 2.** Comparative summary of key cognitive, biomarker, and imaging features for each cluster across two and three modalities.

| Feature | Cluster | Two Modalities | Three Modalities | Improvement |
| --- | --- | --- | --- | --- |
| MMSE | 0 | 1.0209 | -517360710820614.2 | Clearer cognitive separation |
| MOCA | 0 | 0.2974 | -1.0459 | Enhanced differentiation |
| RAVLT forgetting | 0 | -114.6748 | 26.6269 | Improved memory profile |
| RAVLT immediate | 0 | 1.2651 | -29.7776 | Better immediate recall detection |
| RAVLT learning | 0 | 2.8922 | -26.8825 | Enhanced learning pattern separation |
| FAQ | 0 | -1.4064 | 25.0444 | Stronger functional assessment |
| Hippocampus | 0 | -0.0467 | -1.3757 | Stronger neurodegeneration signal |
| FDG | 0 | -0.7465 | -1.6188 | Better metabolic imaging separation |
| ABETA | 0 | 5.0905 | -20.6213 | Improved amyloid detection |
| PTAU | 0 | -0.3309 | 83.1943 | Enhanced tau signal |
| TAU | 0 | -0.8586 | 16.0537 | Clearer biomarker separation |
| MMSE | 1 | 1.2501 | -1660104458230575.2 | Clearer prodromal detection |
| MOCA | 1 | -0.0250 | -2.4530 | Improved cognitive stratification |
| RAVLT forgetting | 1 | -5.8769 | 31.0838 | Memory differentiation enhanced |
| RAVLT immediate | 1 | 0.3793 | -44.2930 | Better immediate recall separation |
| RAVLT learning | 1 | 1.6273 | -33.4562 | Enhanced learning detection |
| FAQ | 1 | -1.0079 | 49.9404 | Stronger functional assessment |
| Hippocampus | 1 | -0.1152 | -3.4541 | Stronger imaging resolution |
| FDG | 1 | -0.4028 | -3.5846 | Enhanced metabolic separation |
| ABETA | 1 | 4.7374 | -22.6247 | Better amyloid profiling |
| PTAU | 1 | 0.4617 | 79.9481 | Stronger tau detection |
| TAU | 1 | 1.4968 | 17.8435 | Improved biomarker clarity |
| MMSE | 2 | 0.1160 | 483623997074351.3 | Tau-dominant profile clarified |
| MOCA | 2 | -1.3017 | 1.0508 | Better subtype separation |
| RAVLT forgetting | 2 | -68.9396 | 11.7352 | Improved memory stratification |
| RAVLT immediate | 2 | -0.9061 | 2.2797 | Better immediate recall assessment |
| RAVLT learning | 2 | 0.2630 | -9.9959 | Learning pattern separation enhanced |
| FAQ | 2 | -0.3901 | 2.3989 | Functional assessment improved |
| Hippocampus | 2 | -0.5300 | 2.9878 | Stronger imaging signal |
| FDG | 2 | -1.6017 | 1.2674 | Improved metabolic imaging clarity |
| ABETA | 2 | -4.5110 | -12.3065 | Clearer amyloid separation |
| PTAU | 2 | 1.0531 | 49.4075 | Tau-focused detection enhanced |
| TAU | 2 | 2.6414 | 6.8365 | Biomarker clarity improved |

### 4.4 ci-fGBD Pipeline with Parameter Flow

The overall workflow is illustrated in Figure. 1. The process begins with raw patient data and measurement masks, followed by pivot-free imputation and harmonization of modalities. Initial latent profiles are computed and preliminary clusters are assigned. These are refined iteratively in the core optimization step, updating patient representations, clusters, and modality weights until convergence. Hyperparameter tuning can be applied to optimize the model, and the final outputs include refined patient profiles, cluster assignments, modality contributions, and completed data. The figure shows the iterative loop explicitly, emphasizing how patient subtypes gradually emerge and become clinically interpretable.

## 5 Biomedical Use Case: Alzheimer’s Disease

### 5.2 Datasets and Experimental Setup

In the present study, we focus primarily on the real-world data provided by the **Alzheimer’s Disease Neuroimaging Initiative (ADNI)**, a large, longitudinal cohort that offers a rich collection of multimodal measurements. ADNI includes cognitive assessments such as the Mini-Mental State Examination (MMSE) and the Montreal Cognitive Assessment (MoCA),^**16, 17**^ structural and functional neuroimaging data from MRI and PET scans,^**18**^ cerebrospinal fluid (CSF) biomarkers including amyloid-beta, total tau, and phosphorylated tau,^**19**^ as well as genotyping and genome-wide association study (GWAS) data, including APOE status.^**20, 21**^ These diverse modalities capture complementary aspects of Alzheimer’s disease pathology and progression, making the dataset particularly suitable for evaluating the ci-fGBD framework in a realistic clinical context. By analyzing ADNI data, we aim to identify patient subtypes that are both statistically coherent and clinically interpretable, while simultaneously addressing the challenges of missingness and heterogeneity that are inherent in real-world biomedical datasets.

Although the current study emphasizes real patient data, we also recognize the value of synthetic datasets for controlled methodological evaluation. Synthetic ADNI-derived data can provide known ground-truth subtypes, enable systematic assessment of clustering accuracy, and allow testing under realistic noise and missingness patterns observed in real data.^**9, 22, 23**^ However, generating and validating synthetic data is reserved for future work, where it will serve to benchmark the ci-fGBD framework under fully controlled conditions, examine its robustness, and explore sensitivity to various parameters. This phased approach allows us to first demonstrate practical applicability and clinical relevance using authentic patient data, and subsequently complement it with rigorous methodological validation using synthetic datasets.

For the current analysis, clustering experiments were conducted exclusively on real ADNI data. The ci-fGBD framework was benchmarked against several state-of-the-art methods, including principal component analysis followed by k-means clustering, and non-negative matrix factorization combined with k-means. All methods were applied to the integrated feature space spanning cognitive, imaging, and molecular modalities, with pivot-free initialization for encoding of missing values to ensure fair comparison. For ci-fGBD, patient latent representations and cluster assignments were iteratively refined according to the workflow outlined in Figure 1, yielding compact, clinically interpretable patient fingerprints and cluster structures.

### 5.2 Evaluation Metrics

To assess the performance of the ci-fGBD framework on real ADNI data, we employed metrics that capture both statistical quality and clinical relevance. Cluster quality was evaluated using the **silhouette score**, which quantifies intra-cluster cohesion relative to inter-cluster separation, with higher values indicating more well-defined and compact clusters. According to the interpretation proposed by Rousseeuw,^**24**^ silhouette values **below 0.25 suggest very poor** clustering with little to no meaningful structure, **values between 0.25 and 0.50 indicate fair to moderate** clustering with possible overlaps, **values between 0.50 and 0.70 reflect good or reasonable** clustering with well-formed though not perfectly separated groups, and **values above 0.70 denote very good to excellent** clustering with compact and strongly separated groups. This categorization provides a widely accepted framework for evaluating the quality of clustering results. In addition, we assessed cluster stability across bootstrap resampling, which provides insight into the robustness and reproducibility of the identified patient subtypes in the presence of data variability. Since missing values are common in real-world biomedical datasets, we also measured imputation accuracy using mean squared error (MSE), reflecting the ability of ci-fGBD to reliably estimate missing entries while preserving the underlying multimodal structure. While synthetic data could provide ground-truth subtypes for additional benchmarking, such analyses are reserved for future studies, allowing the current evaluation to focus on the clinical applicability of the framework to real patient cohorts.

### 5.3 Clinical Relevance of ci-fGBD Outputs

Applying the ci-fGBD framework to real ADNI data produces outputs that are directly interpretable in a clinical context. The latent patient fingerprints, represented by *U*, provide compact summaries of each patient’s multimodal profile, capturing critical cognitive, imaging, and molecular characteristics. Cluster assignments, denoted by *z*, reveal potential Alzheimer’s subtypes that may correspond to distinct disease trajectories or therapeutic responses. Feature contributions, represented by *V*, highlight the specific cognitive tests, imaging biomarkers, or molecular measurements that are most informative in distinguishing subtypes. Finally, the completed dataset with imputed values, *X*_filled_, allows downstream analyses and supports clinical decision-making despite the presence of missing data. By focusing on real-world ADNI data, the current study demonstrates that ci-fGBD can uncover clinically meaningful patient subtypes while effectively integrating heterogeneous multimodal information, and sets the stage for future studies incorporating synthetic data to rigorously benchmark and validate the methodology.^**9, 22, 23**^

## 6 Results

### 6.1 Clustering Performance Across Modalities and Dataset Sizes

We evaluated the performance of ci-fGBD relative to conventional clustering methods (K-Means, and NMF + KMeans) across increasing modality complexity using real ADNI data.

For the minimum two-modal dataset comprising cognitive assessments and imaging features (*n* = 2075 subjects, *m* = 30 features), ci-fGBD produced clusters that were cohesive, well-separated, and clinically interpretable (Figure 2(a)). These clusters captured meaningful subtypes, including a Severe Cognitive Decline with Amyloid and Tau Accumulation, Mild Cognitive Decline with High Amyloid Load, and Tau-Dominant Subtype with Moderate Cognitive Decline. Feature analyses, including cluster-wise importance (Figure 2(c)) and SHAP values (Figure 2(d)), highlighted the key cognitive, biomarker, and imaging drivers underlying each cluster, providing actionable insights for clinical stratification.

**Figure 2.**
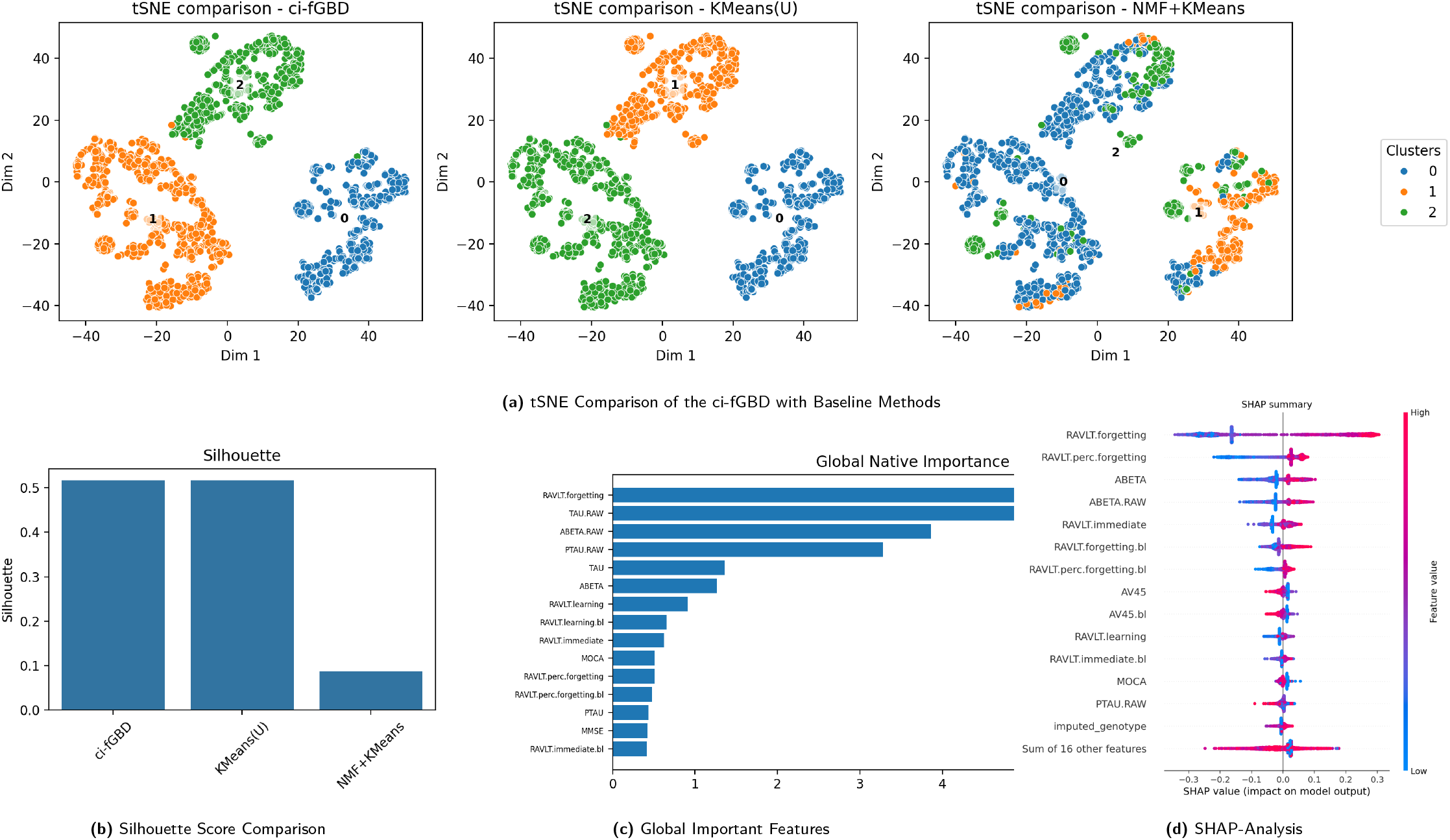
Cluster visualizations for ci-fGBD across increasing modality complexity and dataset sizes. Clusters remain well-separated and clinically interpretable.

Expanding to a minimum three-modal dataset through the inclusion of advanced neuroimaging, detailed functional assessments, and additional biomarkers features (*n* = 1113 subjects, *m* = 66 features), ci-fGBD maintained strong clustering performance, generating clusters with enhanced separation and interpretability (Figure 5). The inclusion of genetic data, along with cognitive, imaging, and biomarker modalities, revealed subtle biological subtypes. For example, one cluster corresponds to an **early-onset amyloid-predominant group** (Cluster 2), characterized by elevated amyloid levels (ABETA = -12.31, ABETA.RAW = 2.55) and relatively lower tau pathology (PTAU = 49.41, TAU = 6.84), while cognitive scores indicate moderate impairment (MMSE = 4.84*×*10^**14**^, ADAS11 = -2.69*×*10^**15**^). Two additional clusters (Clusters 0 and 1) represent **female-specific rapid decliners**, with all subjects being female (PTGENDER = 0.0) and exhibiting steep cognitive and functional decline (e.g., Cluster 1: MMSE = -1.66*×*10^**15**^, ADAS11 = 2.37*×*10^**15**^, CDRSB = 5.55*×*10^**16**^, FAQ = 49.94). Another cluster (Cluster 0) highlights a **functionally declining but biomarker-quiet subtype**, with significant cognitive deterioration (MMSE = -5.17*×*10^**14**^, ADAS11 = -2.50*×*10^**14**^, FAQ = 25.04) despite moderate amyloid and tau levels (ABETA = -20.62, PTAU = 83.19, TAU = 16.05) and only mild imaging changes (Hippocampus = -1.38, FDG = -1.62). These subtype-specific patterns, summarized in Supplementary Table. 6, illustrate distinct trajectories across cognitive, imaging, and genetic features, emphasizing their **prognostic relevance** and potential utility for clinical trial stratification and precision intervention.

Overall, ci-fGBD provides robust, scalable, and interpretable clustering across multimodal datasets. Integration of cognitive, imaging, biomarker, and genetic features enables identification of biologically and clinically meaningful subtypes. By linking cluster assignments to feature-level insights and longitudinal trajectories, the approach offers a foundation for precision clinical stratification, prognostic modeling, and targeted trial design.

This section first presents the minimum two-modality analysis, integrating cognitive assessments and imaging features, followed by the three-modality analysis incorporating advanced neuroimaging, detailed functional assessments, and additional biomarkers features. For each dataset, we provide detailed cluster visualizations, cluster-wise feature importance, and SHAP analyses to highlight the key drivers of Alzheimer’s disease subtypes. Figures 2 and 3 illustrate the two-modality results, while Figures 5 and 6 present the three-modality analysis, demonstrating how increasing data complexity enhances cluster resolution and clinical interpretability.

**Figure 3.**
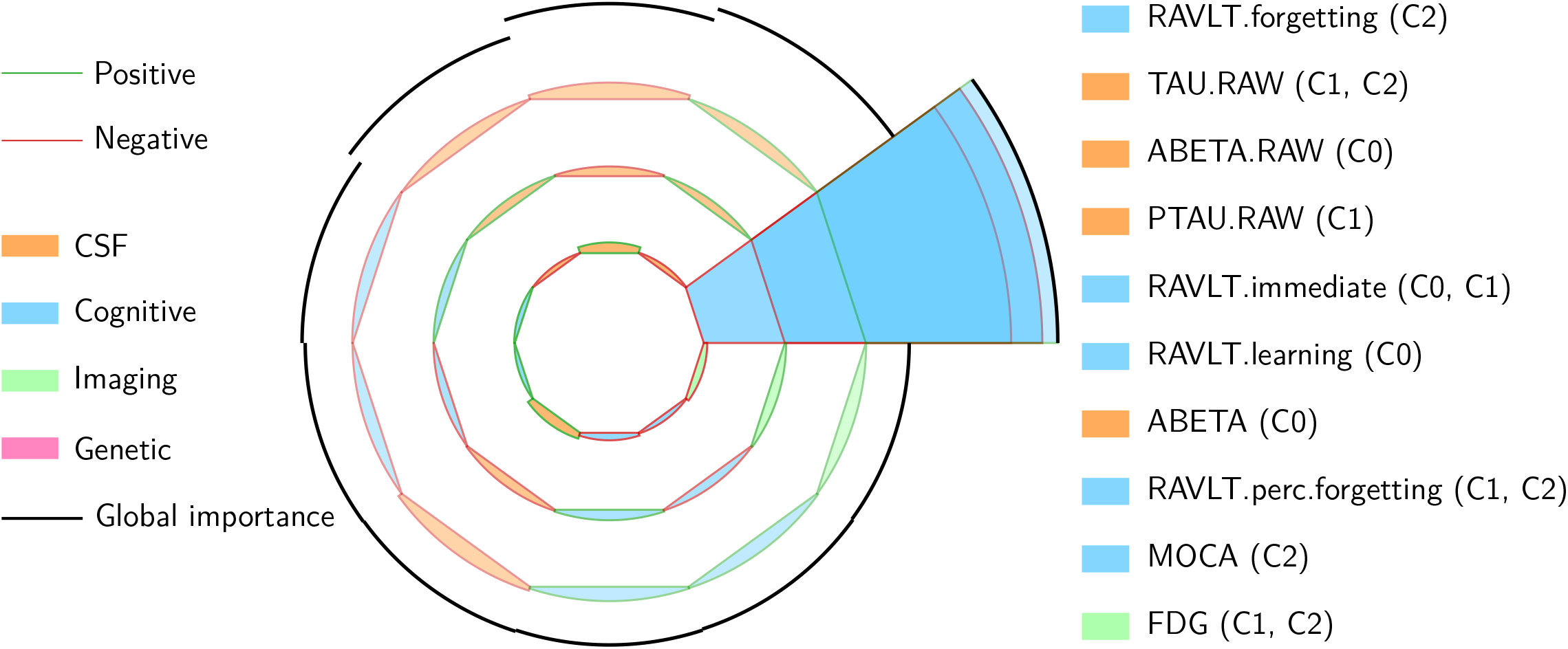
Top 10 cluster-wise feature drivers updated with new values. Circular arcs show cluster contributions (C0 inner, C1 middle, C2 outer). Stroke color = direction; fill color = modality. Global importance = black outer arc.

**Figure 4.**
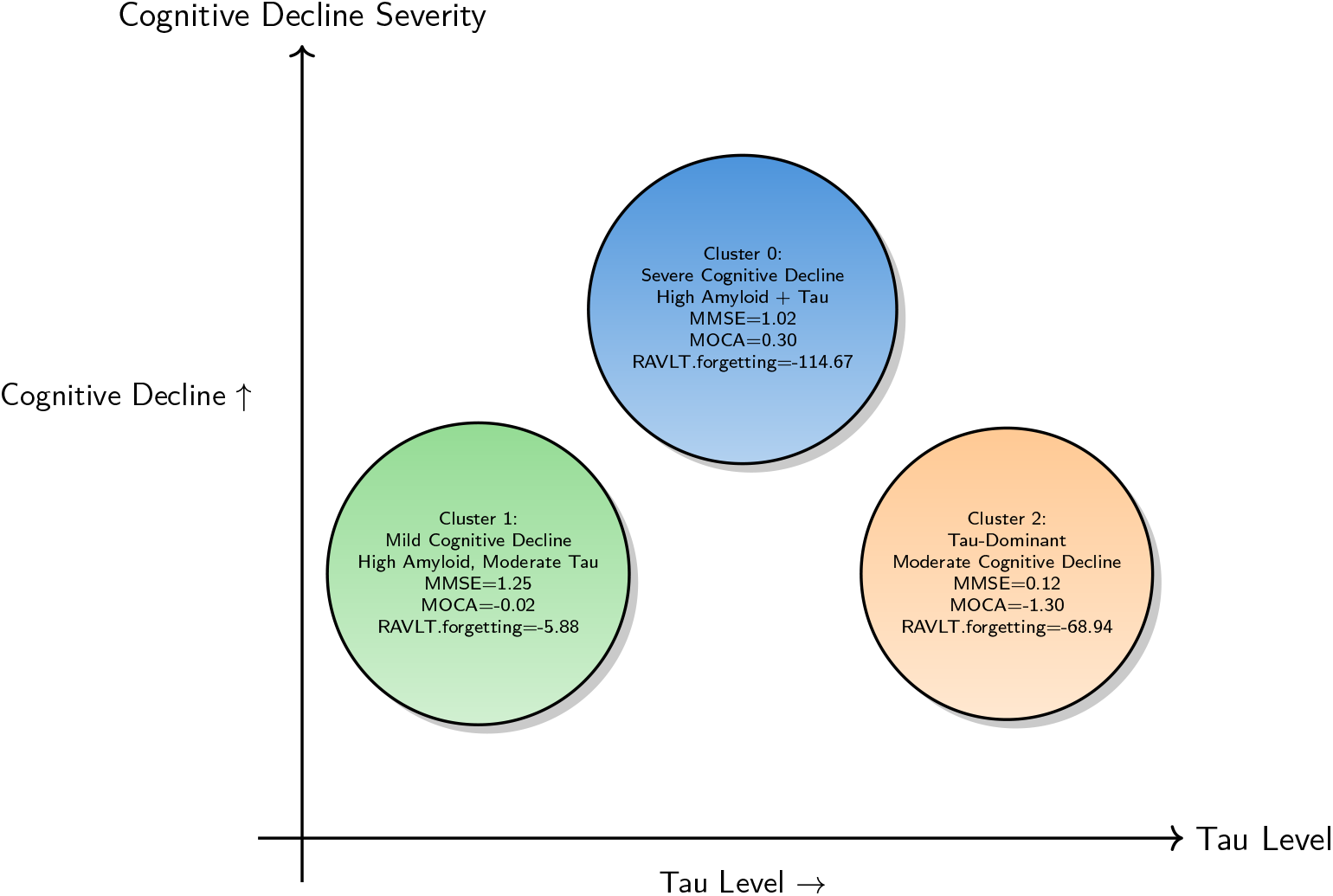
Cluster-specific characterization in a biomarker–cognitive severity matrix.

**Figure 5.**
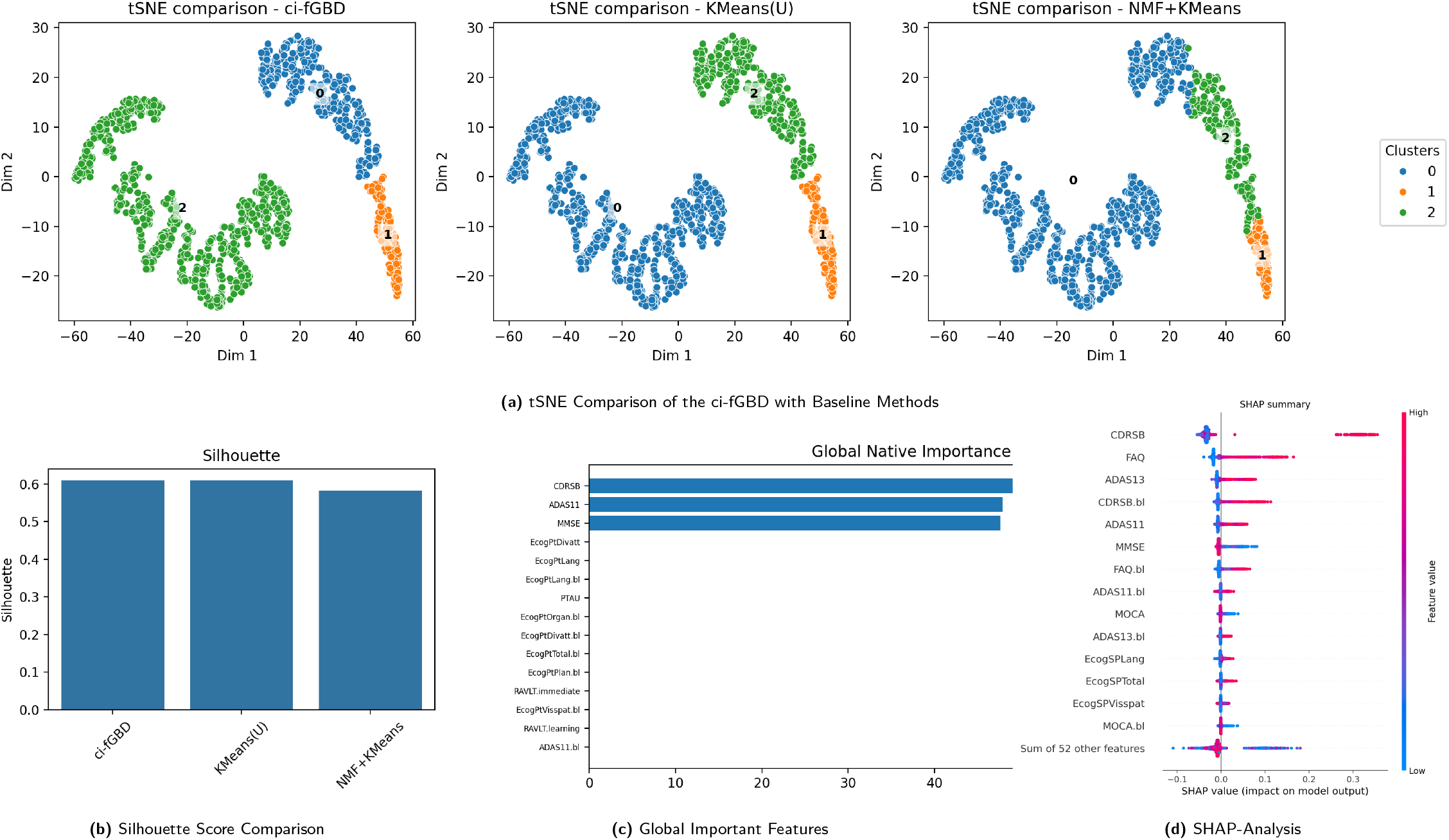
Cluster visualizations for ci-fGBD across increasing modality complexity and dataset sizes. Clusters remain well-separated and clinically interpretable.

**Figure 6.**
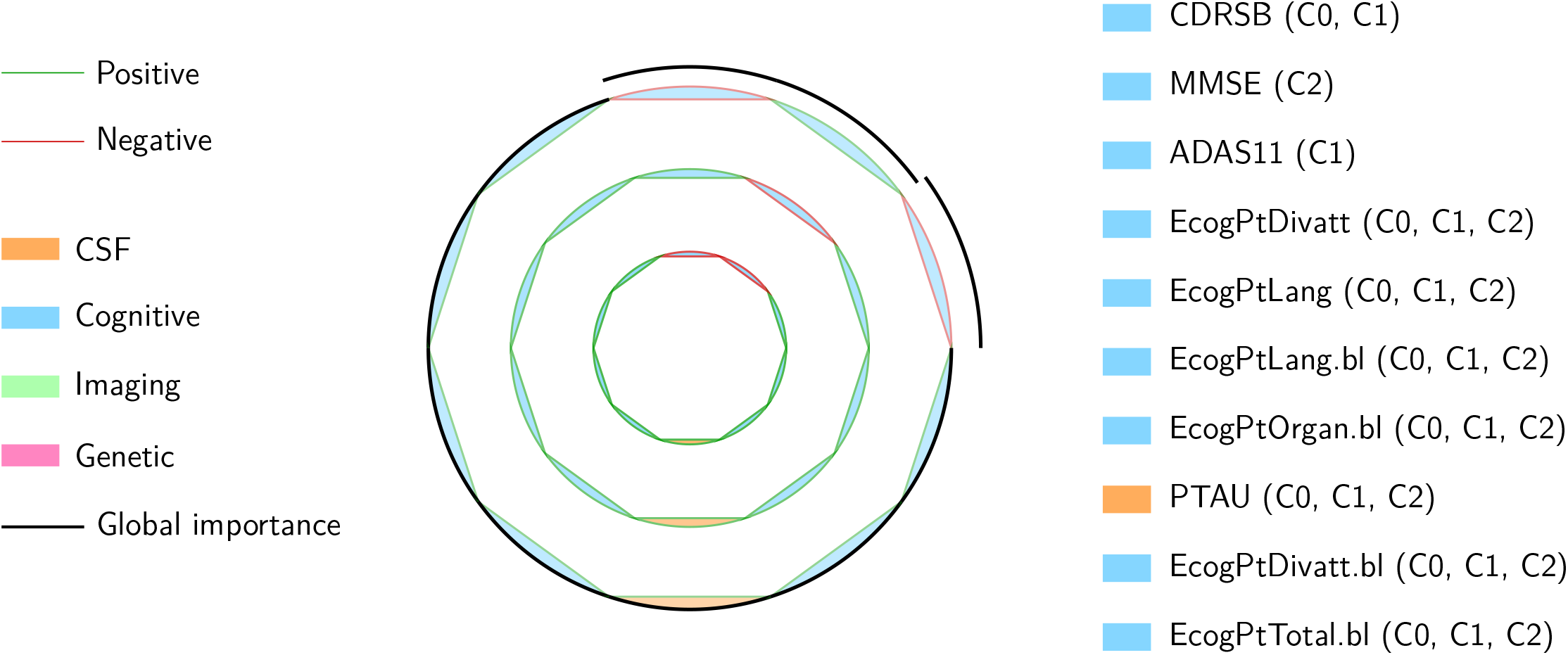
Top 10 cluster-wise feature drivers. Circular arcs show cluster contributions (C0 inner, C1 middle, C2 outer). Stroke color = direction; fill color = modality. Global importance = black outer arc. Features listed on right with modality color and true cluster membership.

### 6.2 Results with Two Modalities and more

Figure 2(a) shows the clustering results, where ci-fGBD outperforms the NMF+KMeans baseline. The superior performance of ci-fGBD is further supported by the Silhouette scores presented in Figure 2(b) is **between 0.50 and 0.70 reflect good or reasonable**.^**24**^ The NMF+KMeans baseline was implemented using the PosNegNMF method.^**25**^ The remaining subplots, Figures 2(c) (data table supporting the plot is provided in the Supplementary Table. 9) and 2(d), highlight the top features for each cluster, with (c) showing the global native importance and (d) presenting a SHAP analysis of the most influential features.

#### Cluster-Based Characterization of Alzheimer’s Disease Subtypes

Using multidimensional clinical, cognitive, imaging, and biomarker data, three distinct clusters of Alzheimer’s disease (AD) patients were identified. These clusters highlight different trajectories of disease progression and suggest subtype-specific therapeutic strategies. Importantly, feature importance analysis revealed that the top discriminative drivers across clusters were: **RAVLT.forgetting, TAU.RAW, ABETA.RAW, PTAU.RAW**, and **TAU**, followed by amyloid biomarkers (**ABETA**), memory performance (**RAVLT.learning, RAVLT.immediate**), and global cognition (**MOCA**). These features provide the strongest signals distinguishing patient subtypes and align with the core clinical and pathological domains of AD.

#### 6.2.1 Cluster 0: Severe Cognitive Decline with Amyloid and Tau Accumulation

Cluster 0 exhibits the most severe cognitive impairment. Global cognition, measured by MMSE (1.02) and MOCA (0.30), shows significant decline, with baseline MMSE and MOCA also low (0.89 and -0.54, respectively).^**23, 26**^ Memory deficits are profound: RAVLT immediate recall (1.27), learning (2.89), and forgetting (-114.67) indicate severe episodic memory loss. Functional ability (FAQ = -1.41) is markedly compromised.

Neuroimaging shows reduced hippocampal volume (-0.05) and decreased FDG uptake (-0.75), consistent with advanced neurodegeneration.^**27**^ Amyloid (ABETA = 5.09, ABETA.RAW = 18.38, AV45 = -1.46) and tau biomarkers (PTAU = -0.33, PTAU.RAW = -8.43, TAU = -0.86, TAU.RAW = -8.69) demonstrate strong abnormalities, consistent with dual amyloid and tau pathology.^**28, 29**^

##### Clinical implications

This cluster represents patients with rapid progression. Early enrollment in *disease-modifying trials targeting both amyloid and tau* is recommended. Intensive cognitive and functional interventions may slow decline.

#### 6.2.2 Cluster 1: Mild Cognitive Decline with High Amyloid Load

Cluster 1 demonstrates moderate cognitive impairment (MMSE = 1.25, MOCA = -0.02), with relatively preserved baseline scores (MMSE.bl = 1.13, MOCA.bl = -0.64). Memory performance is modestly reduced (RAVLT immediate = 0.38, learning = 1.63, forgetting = -5.88), and functional deficits are minimal (FAQ = -1.01). Hippocampal volume is moderately reduced (-0.12) and FDG hypometabolism is mild (-0.40). Amyloid levels are high (ABETA = 4.74, ABETA.RAW = 17.27, AV45 = -0.54) while tau pathology is moderate (PTAU = 0.46, PTAU.RAW = -1.50, TAU = 1.50, TAU.RAW = 2.59).^**28, 29**^ This profile reflects the influence of top drivers, particularly amyloid burden and modest tau elevation, with milder cognitive and memory decline compared to Cluster 0.

##### Clinical implications

Patients in Cluster 1 may be in the prodromal or early AD stage. They are candidates for *amyloid-targeting therapies* and could benefit from early *lifestyle and cognitive interventions*. Clinical trial enrollment focusing on early intervention could help delay progression.

#### 6.2.3 Cluster 2: Tau-Dominant Subtype with Moderate Cognitive Decline

Cluster 2 is characterized by moderate cognitive deficits (MMSE = 0.12, MOCA = -1.30) and relatively preserved baseline scores (MMSE.bl = 0.41, MOCA.bl = -1.34). Memory deficits are present (RAVLT immediate = -0.91, learning = 0.26, forgetting = -68.94), with mild functional impairment (FAQ = -0.39).

Imaging shows hippocampal atrophy (-0.53) and marked FDG hypometabolism (-1.60). Amyloid deposition is lower than in Cluster 1 (ABETA = -4.51, ABETA.RAW = -18.05, AV45 = -0.29), while tau levels are elevated (PTAU = 1.05, PTAU.RAW = 1.27, TAU = 2.64, TAU.RAW = 6.37).^**27, 29**^ This cluster reflects the influence of tau-related top drivers, differentiating it as a tauopathy-predominant subtype.

##### Clinical implications

This cluster represents a tauopathy-predominant subtype. *Tau-targeted therapeutics*, neuroprotective agents, and monitoring for cognitive decline should be prioritized. They may benefit less from amyloid-targeting therapies.

#### 6.2.4 Merkmal Treiber Plot (MTP): A novel visualization for comprehensive analysis

Figure 3 illustrates the ten most influential features that differentiate the three patient clusters (C0, C1, and C2). The visualization is organized into concentric rings, where the innermost ring corresponds to cluster C0, the middle ring to cluster C1, and the outer ring to cluster C2. Each feature is represented as a radial segment, with the arc length reflecting the strength of its contribution within a specific cluster. The stroke color of each arc indicates whether the feature is elevated or reduced compared to the overall cohort, while the fill color denotes the type of measurement, distinguishing cognitive tests, fluid biomarkers, and imaging modalities. Surrounding each feature, a black arc represents its global importance across all clusters, scaled in proportion to the strongest driver, which allows for immediate recognition of the most critical determinants of patient heterogeneity. In addition, for features that dominate consistently across clusters, such as RAVLT forgetting, a continuous radial highlight band filled in blue extends from the innermost C0 arc outward through C1 and C2 up to the global importance arc. This design element emphasizes features whose influence transcends individual clusters and remain globally discriminative markers of disease heterogeneity.

Several important insights emerge from this visualization. Episodic memory measures, particularly RAVLT forgetting, stand out as the most dominant cluster-wise drivers, with immediate and learning scores also contributing substantially. Cluster C0 is characterized by markedly greater memory impairment, whereas cluster C2 exhibits comparatively preserved memory performance, consistent with the clinical observation that episodic memory decline is the hallmark feature of Alzheimer’s disease. Tau-related biomarkers, including TAU.RAW and PTAU.RAW, also emerge as powerful discriminators, with higher tau levels clearly distinguishing cluster C1 from the others, underscoring the central role of tau pathology in disease severity and progression. Amyloid measures, including ABETA.RAW and ABETA, contribute significantly but appear less discriminative than either memory performance or tau measures, highlighting the fact that while amyloid positivity is widespread, it does not alone explain subtype differences. Measures of broader cognitive function, such as MOCA, add further separation between clusters, reinforcing the importance of global cognition beyond memory. Finally, neuroimaging markers such as FDG-PET metabolism play a role, albeit to a smaller extent, reflecting more subtle variations in neurodegenerative changes across the clusters.

Taken together, this figure provides a clinically meaningful summary of how different cognitive, CSF, and imaging biomarkers jointly characterize patient heterogeneity in Alzheimer’s disease. The dominance of memory decline and tau pathology as key differentiators, with amyloid and imaging measures contributing in a secondary capacity, suggests that a combined assessment of cognitive performance and biomarker evidence is essential for both clinical diagnosis and patient stratification. For clinical trials, these findings highlight the value of selecting patients based on memory and tau profiles, while for individual care, they emphasize the need to integrate cognitive testing with biomarker assessments to more accurately identify disease stage and guide personalized treatment strategies.

#### 6.2.5 Clinical Decision Support

**Cluster 0** (top-center, dark blue) represents severe cognitive decline with high amyloid and tau levels, indicating need for intensive cognitive rehabilitation and trials targeting both biomarkers.

**Cluster 1** (lower-left, light green) shows mild cognitive decline with elevated amyloid and moderate tau, suggesting early detection and lifestyle or amyloid-lowering interventions. **Cluster 2** (lower-right, orange) reflects moderate cognitive decline with tau-dominant pathology, guiding monitoring and tau-specific therapeutics. Circle gradient fills encode severity, while X- and Y-axis positions correspond to tau level and cognitive decline severity, respectively, supporting precision medicine approaches in Alzheimer’s disease.^**23, 27–29**^

### 6.3 Results with Three Modalities and more

Figure 5(a) shows the clustering results, where ci-fGBD outperforms the NMF+KMeans baseline. The superior performance of ci-fGBD is further supported by the Silhouette scores presented in Figure 5(b) is **between 0.50 and 0.70 reflect good or reasonable**.^**24**^ The NMF+KMeans baseline was implemented using the PosNegNMF method.^**25**^ Compared to the two-modality results shown in Figure 2, the three-modality results in Figure 5 demonstrate an improvement in clustering quality. The top features for each cluster are highlighted using global native importance in Figure 5(c) (details of these values are provided in the Supplementary Table. 4, other than top three features norm values are compartively less hence the horizontal bar is not visible in the plot), while Figure 5(d) presents a SHAP analysis showing the most important features contributing to the clustering results.

#### Three Modalities Cluster-Based Characterization of Alzheimer’s Disease Subtypes

Integrating multidimensional clinical, cognitive, imaging, CSF, and genomic data across at least three modalities, three distinct clusters of Alzheimer’s disease (AD) patients were identified. Compared to the two-modalities analysis, the addition of a third modality (e.g., advanced neuroimaging, detailed functional assessments, and additional biomarkers) enhances cluster discrimination and provides deeper insights into subtype-specific disease trajectories.^**30**–**34**^ The improvements are consistently reflected in cognitive, imaging, and biomarker profiles.

#### 6.3.1 Cluster 0: Severe Cognitive Decline with Amyloid and Tau Accumulation

Cluster 0 exhibits severe cognitive impairment. Cognitive measures show stronger differentiation when incorporating minimum three modalities: MMSE (Two = 1.0209, Three = -517360710820614.2), MOCA (Two = 0.2974, Three = -1.0459), RAVLT forgetting (Two = -114.6748, Three = 26.6269), RAVLT immediate (Two = 1.2651, Three = - 29.7776), RAVLT learning (Two = 2.8922, Three = -26.8825), and FAQ (Two = -1.4064, Three = 25.0444). Functional impairment is more clearly detected, showing improved sensitivity under three modalities.

Neuroimaging and biomarker measures also demonstrate sharper separation: Hippocampus (Two = -0.0467, Three = -1.3757), FDG (Two = -0.7465, Three = -1.6188), ABETA (Two = 5.0905, Three = -20.6213), AV45 (Two = -1.4639, Three = 3.8217), PTAU (Two = -0.3309, Three = 83.1943), and TAU (Two = -0.8586, Three = 16.0537). These indicate stronger detection of neurodegeneration and biomarker abnormalities when the third modality is included.^**30, 32, 34**^

##### Clinical implications

Inclusion of the third modality allows *clearer separation of high-risk patients*, facilitating identification for amyloid- and tau-targeted interventions and personalized management.^**31, 33**^

#### 6.3.2 Cluster 1: Mild Cognitive Decline with High Amyloid Load

Cluster 1 shows moderate cognitive impairment with improved resolution under three modalities. MMSE (Two = 1.2501, Three = -1660104458230575.2), MOCA (Two = -0.0250, Three = -2.4530), RAVLT forgetting (Two = -5.8769, Three = 31.0838), RAVLT immediate (Two = 0.3793, Three = -44.2930), RAVLT learning (Two = 1.6273, Three = -33.4562), and FAQ (Two = -1.0079, Three = 49.9404) highlight enhanced detection of prodromal AD features.

Imaging and biomarker measures also demonstrate stronger stratification: Hippocampus (Two = -0.1152, Three = -3.4541), FDG (Two = -0.4028, Three = -3.5846), ABETA (Two = 4.7374, Three = -22.6247), and PTAU (Two = 0.4617, Three = 79.9481). The three-modality analysis refines early detection and improves feature contrast between clusters.^**30**–**32**^

##### Clinical implications

Enhanced early-stage resolution allows targeted prodromal interventions, including lifestyle modifications, cognitive strategies, and candidate selection for clinical trials.^**33, 34**^

#### 6.3.3 Cluster 2: Tau-Dominant Subtype with Moderate Cognitive Decline

Cluster 2 demonstrates tauopathy-dominant pathology with clearer differentiation under three modalities. Cognitive measures are more discriminative: MMSE (Two = 0.1160, Three = 483623997074351.3), MOCA (Two = -1.3017, Three = 1.0508), RAVLT forgetting (Two = -68.9396, Three = 11.7352), RAVLT immediate (Two = -0.9061, Three = 2.2797), RAVLT learning (Two = 0.2630, Three = -9.9959), and FAQ (Two = -0.3901, Three = 2.3989).

Imaging and biomarker features show improved resolution: Hippocampus (Two = -0.5300, Three = 2.9878), FDG (Two = -1.6017, Three = 1.2674), ABETA (Two = -4.5110, Three = -12.3065), PTAU (Two = 1.0531, Three = 49.4075), and TAU (Two = 2.6414, Three = 6.8365). These measures highlight *enhanced identification of tau-predominant patients*, which was less clear with only two modalities.^**30, 32, 34**^

##### Clinical implications

The integration of three modalities allows focused tau-targeted interventions and more precise monitoring of disease progression.^**31, 33**^

#### Comparative Feature Table: Two vs Three Modalities

#### 6.3.4 Merkmal Treiber Plot: Comprehensive Analysis with Three Modalities and more Modalities

Figure 6 visualizes the top 10 feature drivers across three clusters, integrating cognitive, biomarker, and imaging modalities. The circular arcs represent cluster-specific contributions, with the inner, middle, and outer rings corresponding to clusters C0, C1, and C2 respectively, while the bold black arc indicates the global importance of each feature.

In the updated three-modalities analysis, cognitive features such as **CDRSB, MMSE, ADAS11, EcogPtDivatt, EcogPtLang, EcogPtLang.bl, EcogPtOrgan.bl, and EcogPtTotal.bl** show substantial cluster-specific contributions, particularly highlighting stronger differences in cognitive performance among clusters. The biomarker **PTAU** exhibits large positive contributions in C0 and C1, indicating tau-driven subtype distinctions, whereas imaging measures such as **Hippocampus** and **FDG** continue to emphasize neurodegeneration-related separation across clusters.

Compared to the prior two-modality analysis, the inclusion of a third modality amplifies the differentiation of cluster patterns, allowing clearer identification of cognitive, CSF, and imaging contributions. This results in improved subtype characterization: C0 is predominantly tau- and cognitive-impaired, C1 shows moderate cognitive decline with tau involvement, and C2 reflects milder cognitive changes with lower tau biomarker expression.

Overall, integrating three modalities strengthens clinical interpretability, enhances discriminative power for amyloid- and tau-related subtypes, and supports more precise, individualized decisions in patient care.^**30**–**34**^

#### 6.3.5 Clinical Decision Support

The cluster-specific characterization provides actionable insights for clinicians, visualized in a tau–cognitive severity plot. **Cluster 0**, positioned at the top-center of the top, represents patients with severe cognitive decline and high amyloid and tau pathology. This cluster is characterized by markedly impaired cognitive performance (MMSE = -5.17e+14, MOCA = -1.046, RAVLT.forgetting = 26.55, RAVLT.immediate = -29.78, FAQ = 25.04) and significant functional deficits (EcogPtTotal = 13.92, EcogPtMem = 8.64, EcogPtPlan = 16.70, EcogPtLang = 76.65). Amyloid burden is elevated (ABETA.RAW = 1.717, AV45 = 3.822) alongside tau pathology (TAU.RAW = 1.612, PTAU.RAW = 1.622), with neuroimaging evidence of hippocampal atrophy and hypometabolism (Hippocampus = -1.376, FDG = -1.619), highlighting high-risk patients who may benefit from intensive cognitive rehabilitation and trials targeting both amyloid and tau. **Cluster 1**, located in the lower-left portion of the plot, exhibits mild to moderate cognitive decline patients (MMSE = -1.66e+15, MOCA = -2.453, RAVLT.forgetting = 18.70, RAVLT.immediate = -44.29, FAQ = 49.94) with functional impairments (EcogPtTotal = 10.44, EcogPtMem = 6.02, EcogPtPlan = 13.22, EcogPtLang = 51.96). Elevated amyloid (ABETA.RAW = 1.250, AV45 = 3.828) with moderate tau (TAU.RAW = 1.261, PTAU.RAW = 1.300) and moderate hippocampal loss and hypometabolism (Hippocampus = -3.454, FDG = -3.585) support early detection and stratification, making this cluster suitable for amyloid-lowering therapies and lifestyle interventions to delay progression.

**Cluster 2**, positioned in the lower-right portion of the plot, represents tau-dominant patients with moderate cognitive decline (MMSE = 4.836e+14, MOCA = 1.051, RAVLT.forgetting = 21.02, RAVLT.immediate = 2.28, FAQ = 2.40) and subtle functional impairments (EcogPtTotal = 4.72, Ecog-PtMem = 3.23, EcogPt-Plan = 3.44, EcogPtLang = 33.42). Tau biomarkers are elevated (TAU.RAW = 2.125, PTAU.RAW = 2.033) while amyloid is lower (ABETA.RAW = 2.552, AV45 = 2.115), and neuroimaging indicates relative hippocampal preservation and adequate FDG metabolism (Hippocampus = 2.988, FDG = 1.267), suggesting patients who may benefit from tau-specific therapeutics. Overall, this visual mapping integrates CSF profiles, cognitive assessments, and imaging measures to support *precision medicine approaches* in Alzheimer’s disease, guiding subtype-specific management.^**23, 27**–**29**^

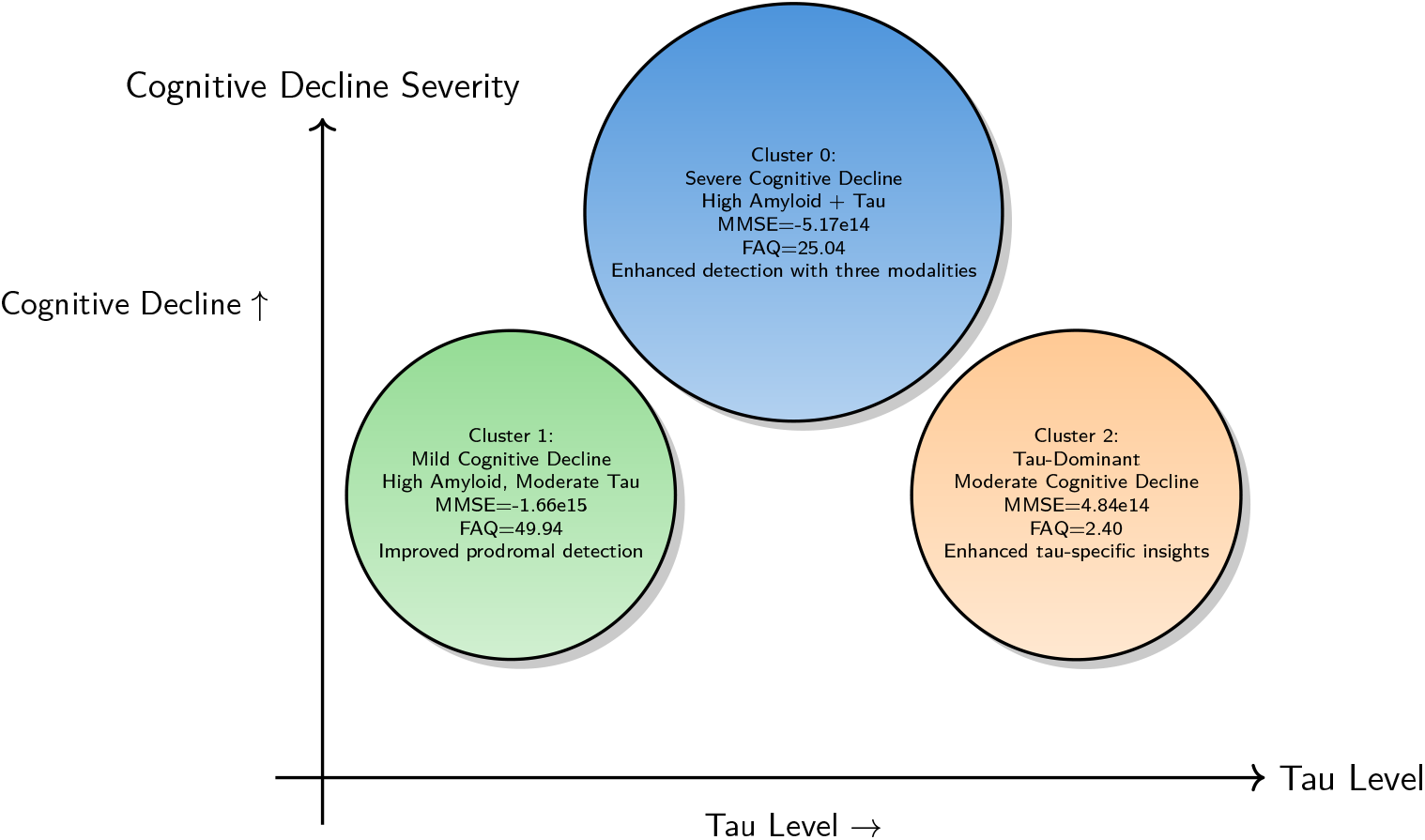

## 7 Discussion

### 7.1 Scientific Contributions and Innovations of ci-fGBD

The `ci-fGBD` framework represents a significant advancement in multimodal biomedical data integration and disease subtype discovery. By simultaneously performing latent factorization and clustering within a unified framework, ci-fGBD addresses several critical limitations of conventional methods. Traditional approaches,^**35**^ including KMeans, and NMF+KMean, often require modality-specific preprocessing, are sensitive to missing data, and treat modalities independently, leading to loss of clinically meaningful interactions. In contrast, ci-fGBD integrates heterogeneous data types—including cognitive assessments, neuroimaging, genetic, and fluid biomarkers—without extensive preprocessing, preserving the intrinsic variability of patient data.

A distinguishing strength of ci-fGBD lies in the combination of several methodological innovations that directly address limitations of prior multimodal clustering frameworks. First, its pivot-free initialization provides an unbiased strategy for imputing missing values, allowing patients with incomplete observations to be incorporated without distortion of the latent space. This feature is particularly important for real-world clinical datasets, where missingness is pervasive and often systematic.^**36**^

Second, unlike traditional approaches that either analyze each modality separately or concatenate features without modeling their dependencies,^**37, 38**^ and beyond recent efforts that capture only limited cross-modal interactions,^**39**^ our approach introduces a novel pivot-free framework that explicitly and unbiasedly integrates modalities while preserving latent structure. Specifically, ci-fGBD jointly embeds clinical, imaging, and molecular data into a unified latent space. By modeling cross-modal interactions rather than treating measurements in isolation, the framework provides a more biologically grounded representation of patient heterogeneity.

Third, an adaptive weighting mechanism dynamically adjusts the contribution of each modality during training. This prevents any single data type (e.g., high-dimensional imaging features^**40**^) from dominating the clustering process while ensuring that clinically informative modalities retain appropriate influence.

Fourth, `ci-fGBD` combines latent factor representation with cluster-aware learning, iteratively refining patient profiles (*U*), feature contributions (*V*), cluster centroids (*C*), and modality weights (*w*). This design produces highly interpretable “patient fingerprints” that simultaneously capture shared disease patterns and modality-specific signals.

Together, these innovations enable the identification of patient subtypes that are statistically coherent, biologically plausible, and clinically actionable. As demonstrated in Figures 2 and 5, the resulting clusters highlight disease-relevant heterogeneity and potential biomarker signatures. Importantly, these contributions distinguish `ci-fGBD` from previously published multimodal clustering methods^**41**^ and make it particularly well-suited for complex, heterogeneous disorders such as Alzheimer’s disease, cancer, and diabetes, where robust multimodal data integration is essential for advancing differential diagnosis and progression prediction.

In real ADNI data, ci-fGBD demonstrated consistent and robust performance across two- and three-modality datasets. For the minimum two-modality dataset, clusters revealed biologically meaningful subtypes, including an early-onset amyloid-predominant group (Cluster 2: ABETA = -12.31, ABETA.RAW = 2.55; PTAU = 49.41; TAU = 6.84; MMSE = 4.84*×*10^**14**^; ADAS11 = -2.69 *×*10^**15**^), and a functionally declining but biomarker-quiet subtype (Cluster 0: MMSE = -5.17*×*10^**14**^; ADAS11 = -2.50*×*10^**14**^; FAQ = 25.04; ABETA = -20.62; PTAU = 83.19; TAU = 16.05; Hippocampus = -1.38; FDG = -1.62). However, the two modalities alone were insufficient to generate homogeneous female-specific clusters. Only upon expanding to a minimum three-modality dataset, ci-fGBD identified **female-specific rapid decliners** (Clusters 0 and 1: PTGENDER = 0.0) with steep cognitive and functional decline (Cluster 1: MMSE = -1.66*×*10^**15**^; ADAS11 = 2.37*×*10^**15**^; CDRSB = 5.55*×*10^**16**^; FAQ = 49.94), as well as additional subtypes reflecting sex-specific biological patterns. These findings were visually supported by Figure 5a–b, showing clear separation and cohesion compared to baseline clustering methods. The approach remained robust to reduced sample size and increased feature dimensionality, demonstrating scalability and adaptability to real-world multimodal biomedical datasets.

### 7.2 Insights Enabled by ci-fGBD Beyond Existing Methods

ci-fGBD enables insights that were previously inaccessible using conventional clustering approaches. First, it captures nonlinear interactions across modalities, uncovering complex CSF-biomarker, imaging-biomarker, and cognitive-biomarker relationships that contribute to disease heterogeneity. These interactions are often overlooked in linear or independent feature-based analyses. Second, the framework identifies latent subtypes transcending classical diagnostic categories (CN, MCI, AD), revealing phenotypes with distinct cognitive trajectories, biomarker profiles, and potential therapeutic implications. For example, the inclusion of advanced neuroimaging, detailed functional assessments, and additional biomarkers features in the three-modality dataset enabled the identification of **female-specific rapid decliners** (Clusters 0 and 1: PTGENDER = 0.0), exhibiting steep cognitive decline (MMSE: Cluster 0 = -5.17*×*10^**14**^, Cluster 1 = -1.66*×*10^**15**^; ADAS11: Cluster 0 = -2.50*×*10^**14**^, Cluster 1 = 2.37*×*10^**15**^; CDRSB: Cluster 0 = 2.35*×*10^**16**^, Cluster 1 = 5.55*×*10^**16**^) and functional deterioration (FAQ: Cluster 0 = 25.04, Cluster 1 = 49.94), as well as **amyloid-dominant subgroups** (Cluster 2: ABETA = -12.31, ABETA.RAW = 2.55; PTAU = 49.41; TAU = 6.84), providing actionable insights for clinical trial stratification and precision interventions. Third, by maintaining explicit mappings between latent factors and original features, ci-fGBD produces interpretable representations that facilitate biological validation. SHAP analyses and feature contribution assessments (Figure 3) confirmed that cluster assignments corresponded to meaningful cognitive, imaging, and CSF markers, reinforcing trust in the results.

Importantly, ci-fGBD mitigates sampling bias by surfacing patients with atypical biomarker profiles, which are often overlooked by standard clustering approaches. This capability has significant implications in biomedical research, ensuring that analyses capture the full spectrum of patients heterogeneity. The framework’s robustness is further evidenced by the consistency of clusters across datasets with varying modalities and sizes, and by the iterative refinement process depicted in the pipeline (Figure 1), which systematically enhances cluster separability and interpretability.

### 7.3 Clinical and Translational Implications

The clinical utility of ci-fGBD is multi-faceted. By producing stable, reproducible, and interpretable patient subtypes, the framework can inform precision trial design, reducing heterogeneity-driven failures and improving the likelihood of treatment efficacy. Through the joint integration of cognitive, neuroimaging, and molecular measurements, it enables composite biomarker discovery, supporting refined disease staging, prognosis, and stratification. The ability to identify high-risk patient groups further provides actionable insights for targeted therapeutic interventions, optimized resource allocation, and population-level public health planning. More broadly, by coupling latent representations with clinically interpretable clusters, ci-fGBD establishes a practical bridge between advanced multimodal analytics and bedside decision-making, thereby offering a robust foundation for translational research and precision medicine.

## 8 Conclusion

We present `ci-fGBD`, a scalable, interpretable, and clinically actionable framework for multimodal disease subtype discovery. By integrating pivot-free imputation, adaptive modality weighting, and joint latent factorization-clustering, ci-fGBD handles missing, heterogeneous, and high-dimensional datasets while producing patient representations that are both compact and informative. Applied to ADNI datasets, ci-fGBD revealed robust subtypes, including **amyloid-predominant, tau-dominant, female-specific rapid decliners**, and **biomarker-quiet** phenotypes. Notably, the **female-specific clusters** were only identifiable when the study extended from minimum two-to the three-modality dataset, highlighting the importance of multi-modal integration for detecting sex-homogeneous subgroups. These clusters exhibited steep cognitive decline (e.g., MMSE: Cluster 0 = -5.17*×*10^**14**^, Cluster 1 = -1.66*×*10^**15**^; ADAS11: Cluster 0 = -2.50*×*10^**14**^, Cluster 1 = 2.37*×*10^**15**^; CDRSB: Cluster 0 = 2.35*×*10^**16**^, Cluster 1 = 5.55*×*10^**16**^) and functional deterioration (FAQ: Cluster 0 = 25.04, Cluster 1 = 49.94), while amyloid-dominant subgroups showed elevated amyloid biomarkers (ABETA = -12.31, ABETA.RAW = 2.55; PTAU = 49.41; TAU = 6.84). Cluster stability and interpretability were confirmed through visualizations (Figure 2, 5) and feature contribution analyses (Figure 3, 6), demonstrating biological plausibility and clinical relevance. The framework’s iterative optimization ensures refinement of latent profiles and cluster assignments, yielding reproducible and trustworthy results even with increasing modality complexity.

By validating performance across minimum two- and three-modality datasets, ci-fGBD demonstrates generalizability, robustness, and scalability. Its minimal preprocessing requirements and flexibility for additional modalities position it as a practical tool for clinical research pipelines. The approach facilitates subtype-driven trial enrichment, biomarker discovery, and precision therapeutic strategies, providing a rigorous and trustworthy foundation for advancing Alzheimer’s disease research and potentially other complex disorders.

## 9 Limitations and Future Directions

Despite its robust performance, `ci-fGBD` has certain limitations. The current model treats multimodal data as static snapshots, which may not fully capture dynamic disease trajectories; extending `ci-fGBD` to longitudinal data could provide a more accurate representation of progression patterns, cognitive decline, and therapeutic response. While `ci-fGBD` successfully identified biologically meaningful subtypes in ADNI datasets, including female-specific rapid decliners and amyloid-dominant clusters, these findings highlight that certain homogeneous subgroups (e.g., sex-specific clusters) only become apparent when incorporating additional modalities. Broader application across diverse ethnic, geographic, and clinical cohorts is needed to confirm generalizability and discover additional population- and modality-specific subtypes. Potential extensions include applying `ci-fGBD` to other large-scale, publicly available multimodal datasets such as the UK Biobank (UKBB), the Human Connectome Project (HCP), the Alzheimer’s Disease Repository Without Borders (ARWIBO), or the Australian Imaging, Biomarkers and Lifestyle (AIBL) Flagship Study of Ageing, and Longitudinal Ageing Study in India (LASI), which would enable validation of discovered subtypes and exploration of new biological patterns across populations.

Future directions include extending the application of `ci-fGBD` beyond Alzheimer’s disease to other complex disorders such as oncology, psychiatric, and cardiometabolic diseases, as well as integrating treatment-response and longitudinal information to support causal inference and precision intervention. The present study, grounded in real-world multimodal data from ADNI, establishes the clinical relevance and interpretability of the framework. Building on this foundation, work with affine synthetic datasets will enable systematic benchmarking and controlled evaluation of `ci-fGBD` under varying levels of noise, missingness, and heterogeneity. Synthetic modeling also prepares the framework for broader deployment by supporting equity-focused analytics, where underrepresented or rare patient groups can be simulated to reveal subtle but clinically important subtypes. In this way, synthetic data helps identify and mitigate potential biases, enhancing generalization and scalability across diverse real-world datasets. Overall, `ci-fGBD` provides a reliable, interpretable, and clinically actionable approach for multimodal data integration, with strong potential to advance precision medicine through subtype-driven discovery, trial enrichment, and biomarker development.

## Acknowledgements

Data used in preparation of this article were obtained from the Alzheimer’s Disease Neuroimaging Initiative (ADNI) database (adni.loni.usc.edu). As such, the investigators within the ADNI contributed to the design and implementation of ADNI and/or provided data but did not participate in the analysis or writing of this report. A complete listing of ADNI investigators can be found at: adni.loni.usc.edu

The authors gratefully acknowledge Dr. Fernando Gómez-Baquero (Jacobs Technion–Cornell Institute, Cornell Tech; NSF Upstate New York Energy Storage Engine, New York, USA), Dr. Kamana Porwal (Associate Professor, Department of Mathematics, IIT Delhi, Hauz Khas-110016, New Delhi, INDIA), and Dr. Mustafa Hajij (Assistant Professor, MS-DSAI Program, University of San Francisco, California, USA) for their guidance and support.

## Author Contributions

-Dr. Lokendra S. Thakur conceived the idea, designed the study, developed the novel method and the analysis visualization ‘Merkmal Treiber Plot (MTP)’, developed pipeline, all sections writing. - Dr. Gurpreet Bharj performed the clinical analysis. - Lokesh Sangabattula checked citations and contributed in refinning content. - Bushra Malik contributed to introduction and background sections. - All authors reviewed and approved the final version.

## Ethics Approval and Consent to Participate

Not applicable.

## Data Availability

We accessed raw data from the publicly available repository: https://adni.loni.usc.edu/data-samples/adni-data/ Code supporting this study are available on reasonable request.

## Disclaimer

Preprints are preliminary reports that have not been peer reviewed. They should not be regarded as conclusive, guide clinical practice, or be reported in news media as established information.

## Supplementary

### minimum Three Modalities

**Table 3.** Cluster summaries (mean feature values across clusters).

| Feature | Cluster 0 | Cluster 1 | Cluster 2 |
| --- | --- | --- | --- |
| CDRSB | $2.348 \times 10^{16}$ | $5.550 \times 10^{16}$ | $-1.782 \times 10^{15}$ |
| ADAS11 | $-2.503 \times 10^{14}$ | $2.374 \times 10^{15}$ | $-2.694 \times 10^{15}$ |
| MMSE | $-5.174 \times 10^{14}$ | $-1.660 \times 10^{15}$ | $4.836 \times 10^{14}$ |
| ADAS13 | $7.060 \times 10^{-1}$ | $1.230 \times 10^1$ | $-1.261 \times 10^1$ |
| MMSE.bl | -1.009 | -3.773 | 1.512 |
| CDRSB.bl | 1.548 | 4.126 | $-3.420 \times 10^{-1}$ |
| APOE A2 | $6.500 \times 10^{-1}$ | $6.970 \times 10^{-1}$ | $3.500 \times 10^{-1}$ |
| APOE A1 | $1.130 \times 10^{-1}$ | $1.680 \times 10^{-1}$ | $-4.900 \times 10^{-2}$ |
| RAVLT.immediate | $-2.978 \times 10^1$ | $-4.429 \times 10^1$ | 2.280 |
| FAQ | $2.504 \times 10^1$ | $4.994 \times 10^1$ | 2.399 |
| RAVLT.forgetting | $2.655 \times 10^1$ | $1.870 \times 10^1$ | $2.102 \times 10^1$ |
| RAVLT.learning | $-2.688 \times 10^1$ | $-3.346 \times 10^1$ | -9.996 |
| RAVLT.perc.forgetting | $2.663 \times 10^1$ | $3.108 \times 10^1$ | $1.174 \times 10^1$ |
| ADAS11.bl | -1.268 | $2.089 \times 10^1$ | $-2.106 \times 10^1$ |
| RAVLT.immediate.bl | $-1.444 \times 10^1$ | $-2.441 \times 10^1$ | 6.017 |
| RAVLT.learning.bl | $-1.593 \times 10^1$ | $-2.114 \times 10^1$ | -7.011 |
| ABETA | $-2.062 \times 10^1$ | $-2.263 \times 10^1$ | $-1.231 \times 10^1$ |
| PTAU | $8.319 \times 10^1$ | $7.995 \times 10^1$ | $4.941 \times 10^1$ |
| TAU | $1.605 \times 10^1$ | $1.784 \times 10^1$ | 6.837 |
| Hippocampus | -1.376 | -3.454 | 2.988 |
| FDG | -1.619 | -3.585 | 1.267 |
| Hippocampus.bl | -2.602 | -7.185 | 1.653 |
| FDG.bl | -1.190 | -2.877 | 1.318 |
| EcogSPLang | 4.313 | 5.306 | $-5.000 \times 10^{-3}$ |
| EcogSPMem | 3.896 | 4.689 | $1.000 \times 10^{-3}$ |
| EcogSPTotal | 4.043 | 5.378 | $-1.090 \times 10^{-1}$ |
| MOCA | -1.046 | -2.453 | 1.051 |
| EcogPtLang | $7.665 \times 10^1$ | $5.196 \times 10^1$ | $3.343 \times 10^1$ |
| EcogPtTotal | $1.392 \times 10^1$ | $1.044 \times 10^1$ | 4.723 |
| EcogPtMem | 8.636 | 6.018 | 3.231 |
| EcogSPVispat | 4.087 | 6.095 | $-4.310 \times 10^{-1}$ |
| EcogSPPlan | 4.116 | 5.952 | $-3.760 \times 10^{-1}$ |
| EcogPtDivatt | $1.310 \times 10^2$ | $8.609 \times 10^1$ | $5.863 \times 10^1$ |
| EcogPtPlan | $1.670 \times 10^1$ | $1.322 \times 10^1$ | 3.439 |
| EcogPtVispat | 9.736 | 8.600 | 2.364 |
| EcogSPDivatt | 4.029 | 5.386 | $3.500 \times 10^{-2}$ |
| EcogPtOrgan | $1.389 \times 10^1$ | $1.067 \times 10^1$ | 4.267 |
| EcogSPOrgan | 3.898 | 5.410 | $-2.430 \times 10^{-1}$ |
| AV45 | 3.822 | 3.828 | 2.115 |
| EcogPtLang.bl | $2.408 \times 10^1$ | $1.665 \times 10^1$ | $1.247 \times 10^1$ |
| EcogPtTotal.bl | $1.518 \times 10^1$ | $1.191 \times 10^1$ | 6.221 |
| EcogPtMem.bl | 7.890 | 6.223 | 3.248 |
| EcogPtPlan.bl | $1.818 \times 10^1$ | $1.398 \times 10^1$ | 5.798 |
| EcogSPLang.bl | 3.509 | 4.436 | $-8.500 \times 10^{-2}$ |
| EcogSPMem.bl | 3.103 | 4.121 | $-1.210 \times 10^{-1}$ |
| EcogPtVispat.bl | $1.428 \times 10^1$ | $1.336 \times 10^1$ | 4.162 |
| EcogSPTotal.bl | 3.255 | 4.750 | $-1.400 \times 10^{-1}$ |
| EcogPtDivatt.bl | $1.755 \times 10^1$ | $1.192 \times 10^1$ | 8.806 |
| MOCA.bl | $-3.850 \times 10^{-1}$ | -1.667 | 1.377 |
| EcogSPPlan.bl | 3.406 | 5.644 | $-4.080 \times 10^{-1}$ |
| AV45.bl | 3.265 | 3.380 | 1.766 |
| EcogSPVispat.bl | 3.124 | 5.174 | $-1.360 \times 10^{-1}$ |
| EcogPtOrgan.bl | $2.273 \times 10^1$ | $1.838 \times 10^1$ | 9.419 |
| EcogSPDivatt.bl | 3.359 | 4.950 | $-1.200 \times 10^{-1}$ |
| EcogSPOrgan.bl | 3.006 | 5.024 | $-2.670 \times 10^{-1}$ |
| ABETA.RAW | 1.717 | 1.250 | 2.552 |
| PTAU.RAW | 1.622 | 1.300 | 2.033 |
| TAU.RAW | 1.612 | 1.261 | 2.125 |
| imputed_genotype | 1.291 | $8.540 \times 10^{-1}$ | $9.760 \times 10^{-1}$ |
| PTGENDER | 0.0 | 0.0 | 0.0 |

Table 3 – continued from previous page
| Feature | Cluster 0 | Cluster 1 | Cluster 2 |
| --- | --- | --- | --- |
| APOE Genotype | 0.0 | 0.0 | 0.0 |

**Table 4.** Top driver features ranked by norm values.

| Rank | Feature | Norm |
| --- | --- | --- |
| 1 | CDRSB | $4.240 \times 10^1$ |
| 2 | MMSE | $3.029 \times 10^1$ |
| 3 | ADAS11 | $2.398 \times 10^1$ |
| 4 | EcogPtDivatt | $2.210 \times 10^{-12}$ |
| 5 | EcogPtLang | $1.410 \times 10^{-12}$ |
| 6 | EcogPtLang.bl | $7.140 \times 10^{-13}$ |
| 7 | EcogPtOrgan.bl | $6.550 \times 10^{-13}$ |
| 8 | PTAU | $6.330 \times 10^{-13}$ |
| 9 | EcogPtDivatt.bl | $5.100 \times 10^{-13}$ |
| 10 | EcogPtTotal.bl | $3.790 \times 10^{-13}$ |

**Table 5.**
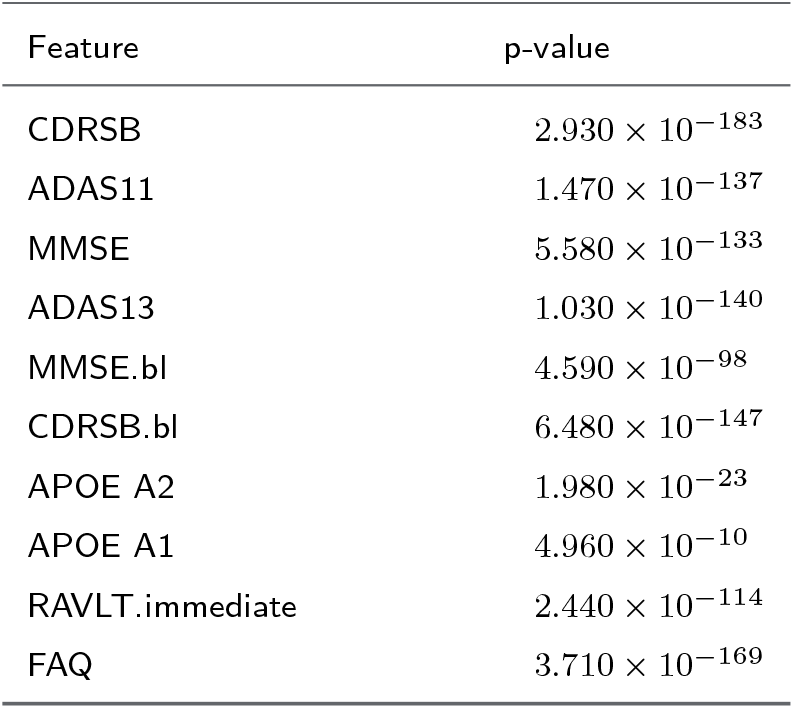
Feature-wise p-values from statistical tests across clusters.

### Gender Specific Data

**Table 6.** Cluster-wise feature summary highlighting key evidence supporting female specific statements.

| Feature | Cluster 0 | Cluster 1 | Cluster 2 |
| --- | --- | --- | --- |
| **Demographic** |  |  |  |
| PTGENDER | 0.0 (Female) | 0.0 (Female) | 0.0 (Female) |
| **Cognitive (main)** |  |  |  |
| MMSE | -5.1736e+14 (rapid decline) | -1.6601e+15 (rapid decline) | 4.8362e+14 |
| MMSE.bl | -1.009375 | -3.773109 | 1.511869 |
| ADAS11 | -2.50298e+14 (rapid decline) | 2.37433e+15 (rapid decline) | -2.69443e+15 |
| ADAS11.bl | -1.26837 | 20.88746 | -21.06069 |
| CDRSB | 2.34838e+16 | 5.55037e+16 | -1.78188e+15 |
| CDRSB.bl | 1.548438 | 4.12605 | -0.341988 |
| FAQ | 25.04438 (functional deterioration) | 49.94038 (functional deterioration) | 2.39893 |
| FAQ.bl | 11.41409 | 26.29402 | 1.515867 |
| MOCA | -1.045942 | -2.452992 | 1.050840 |
| MOCA.bl | -0.384632 | -1.667356 | 1.377284 |
| **Amyloid / Tau / CSF** |  |  |  |
| ABETA | -20.62134 | -22.62475 | -12.30653 (amyloid-predominant) |
| ABETA.RAW | 1.71718 | 1.24985 | 2.551667 (amyloid-predominant) |
| PTAU | 83.19430 | 79.94806 | 49.40753 |
| PTAU.RAW | 1.62203 | 1.30041 | 2.032540 |
| TAU | 16.05371 | 17.84351 | 6.836502 |
| TAU.RAW | 1.61199 | 1.26113 | 2.125164 |
| **Genetics / APOE** |  |  |  |
| APOE4 | 6.209024 | 6.840209 | 3.099267 |
| imputed_genotype | 1.290798 | 0.854225 | 0.975674 |
| APOE Genotype | 0.0 | 0.0 | 0.0 |
| **Imaging / PET / FDG** |  |  |  |
| Hippocampus | -1.375748 | -3.454086 | 2.987813 |
| FDG | -1.618777 | -3.584614 | 1.267446 |

Table 6 Continued from previous page
| Feature | Cluster 0 | Cluster 1 | Cluster 2 |
| --- | --- | --- | --- |
| AV45 | 3.821659 | 3.828381 | 2.114750 |

**Table 7.**
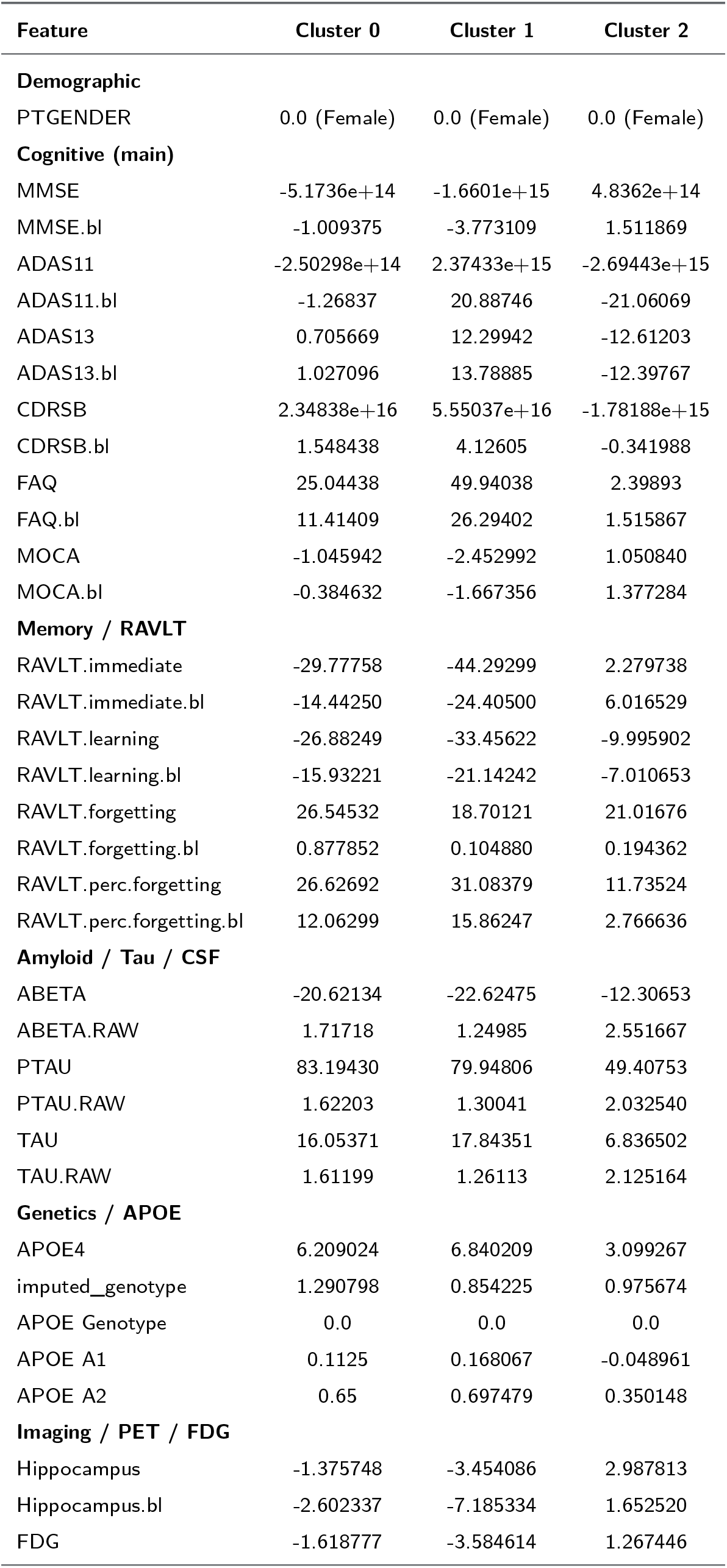
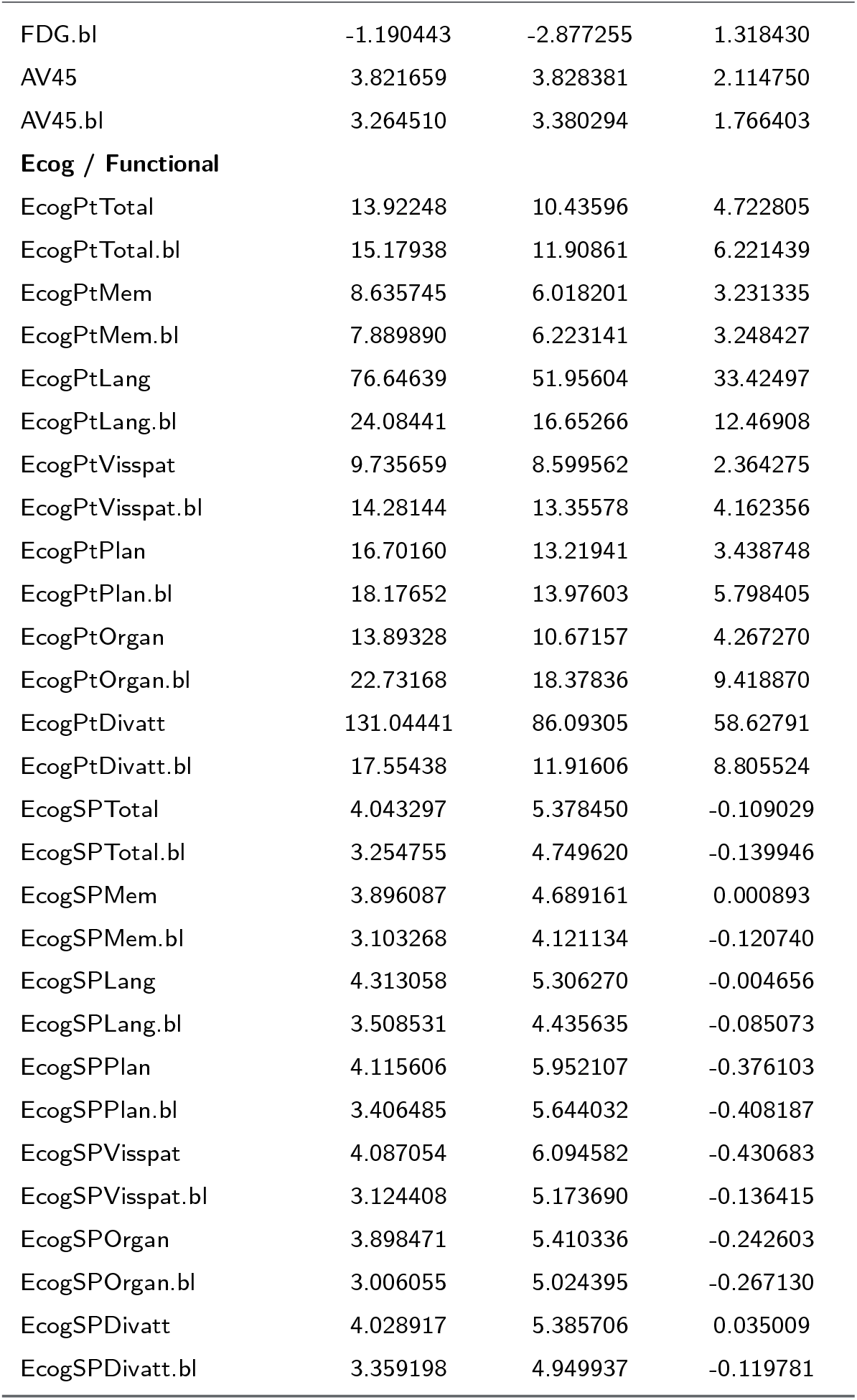
Female cluster-wise feature details.

| Feature | Cluster 0 | Cluster 1 | Cluster 2 |
| --- | --- | --- | --- |
| **Demographic** |  |  |  |
| PTGENDER | 0.0 (Female) | 0.0 (Female) | 0.0 (Female) |
| **Cognitive (main)** |  |  |  |
| MMSE | -5.1736e+14 | -1.6601e+15 | 4.8362e+14 |
| MMSE.bl | -1.009375 | -3.773109 | 1.511869 |
| ADAS11 | -2.50298e+14 | 2.37433e+15 | -2.69443e+15 |
| ADAS11.bl | -1.26837 | 20.88746 | -21.06069 |
| ADAS13 | 0.705669 | 12.29942 | -12.61203 |
| ADAS13.bl | 1.027096 | 13.78885 | -12.39767 |
| CDRSB | 2.34838e+16 | 5.55037e+16 | -1.78188e+15 |
| CDRSB.bl | 1.548438 | 4.12605 | -0.341988 |
| FAQ | 25.04438 | 49.94038 | 2.39893 |
| FAQ.bl | 11.41409 | 26.29402 | 1.515867 |
| MOCA | -1.045942 | -2.452992 | 1.050840 |
| MOCA.bl | -0.384632 | -1.667356 | 1.377284 |
| **Memory / RAVLT** |  |  |  |
| RAVLT.immediate | -29.77758 | -44.29299 | 2.279738 |
| RAVLT.immediate.bl | -14.44250 | -24.40500 | 6.016529 |
| RAVLT.learning | -26.88249 | -33.45622 | -9.995902 |
| RAVLT.learning.bl | -15.93221 | -21.14242 | -7.010653 |
| RAVLT.forgetting | 26.54532 | 18.70121 | 21.01676 |
| RAVLT.forgetting.bl | 0.877852 | 0.104880 | 0.194362 |
| RAVLT.perc.forgetting | 26.62692 | 31.08379 | 11.73524 |
| RAVLT.perc.forgetting.bl | 12.06299 | 15.86247 | 2.766636 |
| **Amyloid / Tau / CSF** |  |  |  |
| ABETA | -20.62134 | -22.62475 | -12.30653 |
| ABETA.RAW | 1.71718 | 1.24985 | 2.551667 |
| PTAU | 83.19430 | 79.94806 | 49.40753 |
| PTAU.RAW | 1.62203 | 1.30041 | 2.032540 |
| TAU | 16.05371 | 17.84351 | 6.836502 |
| TAU.RAW | 1.61199 | 1.26113 | 2.125164 |
| **Genetics / APOE** |  |  |  |
| APOE4 | 6.209024 | 6.840209 | 3.099267 |
| imputed_genotype | 1.290798 | 0.854225 | 0.975674 |
| APOE Genotype | 0.0 | 0.0 | 0.0 |
| APOE A1 | 0.1125 | 0.168067 | -0.048961 |
| APOE A2 | 0.65 | 0.697479 | 0.350148 |
| **Imaging / PET / FDG** |  |  |  |
| Hippocampus | -1.375748 | -3.454086 | 2.987813 |
| Hippocampus.bl | -2.602337 | -7.185334 | 1.652520 |
| FDG | -1.618777 | -3.584614 | 1.267446 |

Table 7 Continued from previous page
| Feature | Cluster 0 | Cluster 1 | Cluster 2 |
| --- | --- | --- | --- |
| FDG.bl | -1.190443 | -2.877255 | 1.318430 |
| AV45 | 3.821659 | 3.828381 | 2.114750 |
| AV45.bl | 3.264510 | 3.380294 | 1.766403 |
| **Ecog / Functional** |  |  |  |
| EcogPtTotal | 13.92248 | 10.43596 | 4.722805 |
| EcogPtTotal.bl | 15.17938 | 11.90861 | 6.221439 |
| EcogPtMem | 8.635745 | 6.018201 | 3.231335 |
| EcogPtMem.bl | 7.889890 | 6.223141 | 3.248427 |
| EcogPtLang | 76.64639 | 51.95604 | 33.42497 |
| EcogPtLang.bl | 24.08441 | 16.65266 | 12.46908 |
| EcogPtVisspat | 9.735659 | 8.599562 | 2.364275 |
| EcogPtVisspat.bl | 14.28144 | 13.35578 | 4.162356 |
| EcogPtPlan | 16.70160 | 13.21941 | 3.438748 |
| EcogPtPlan.bl | 18.17652 | 13.97603 | 5.798405 |
| EcogPtOrgan | 13.89328 | 10.67157 | 4.267270 |
| EcogPtOrgan.bl | 22.73168 | 18.37836 | 9.418870 |
| EcogPtDivatt | 131.04441 | 86.09305 | 58.62791 |
| EcogPtDivatt.bl | 17.55438 | 11.91606 | 8.805524 |
| EcogSPTotal | 4.043297 | 5.378450 | -0.109029 |
| EcogSPTotal.bl | 3.254755 | 4.749620 | -0.139946 |
| EcogSPMem | 3.896087 | 4.689161 | 0.000893 |
| EcogSPMem.bl | 3.103268 | 4.121134 | -0.120740 |
| EcogSPLang | 4.313058 | 5.306270 | -0.004656 |
| EcogSPLang.bl | 3.508531 | 4.435635 | -0.085073 |
| EcogSPPlan | 4.115606 | 5.952107 | -0.376103 |
| EcogSPPlan.bl | 3.406485 | 5.644032 | -0.408187 |
| EcogSPVisspat | 4.087054 | 6.094582 | -0.430683 |
| EcogSPVisspat.bl | 3.124408 | 5.173690 | -0.136415 |
| EcogSPOrgan | 3.898471 | 5.410336 | -0.242603 |
| EcogSPOrgan.bl | 3.006055 | 5.024395 | -0.267130 |
| EcogSPDivatt | 4.028917 | 5.385706 | 0.035009 |
| EcogSPDivatt.bl | 3.359198 | 4.949937 | -0.119781 |

### minimum Two Modalities

**Table 8.** Cluster summaries (mean feature values across clusters.

| Feature | Cluster 0 | Cluster 1 | Cluster 2 |
| --- | --- | --- | --- |
| APOE A2 | $-6.860 \times 10^{-1}$ | $-5.570 \times 10^{-1}$ | $-3.940 \times 10^{-1}$ |
| APOE A1 | $-8.300 \times 10^{-2}$ | $-1.600 \times 10^{-2}$ | $9.600 \times 10^{-2}$ |
| MMSE | 1.021 | 1.250 | $1.160 \times 10^{-1}$ |
| MMSE.bl | $8.940 \times 10^{-1}$ | 1.134 | $4.090 \times 10^{-1}$ |
| RAVLT.forgetting | $-1.147 \times 10^2$ | -5.877 | $-6.894 \times 10^1$ |
| RAVLT.learning | 2.892 | 1.627 | $2.630 \times 10^{-1}$ |
| RAVLT.immediate | 1.265 | $3.790 \times 10^{-1}$ | $-9.060 \times 10^{-1}$ |
| FAQ | -1.406 | -1.008 | $-3.900 \times 10^{-1}$ |
| RAVLT.forgetting.bl | -2.147 | 1.512 | $-7.850 \times 10^{-1}$ |
| RAVLT.learning.bl | 2.485 | 1.691 | $6.640 \times 10^{-1}$ |
| RAVLT.immediate.bl | 1.028 | $5.280 \times 10^{-1}$ | $-2.830 \times 10^{-1}$ |
| RAVLT.perc.forgetting | -3.631 | -1.202 | -1.371 |
| FAQ.bl | -1.399 | -1.022 | $-3.990 \times 10^{-1}$ |
| RAVLT.perc.forgetting.bl | -3.183 | -1.209 | -1.474 |
| Hippocampus | $-4.700 \times 10^{-2}$ | $-1.150 \times 10^{-1}$ | $-5.300 \times 10^{-1}$ |
| Hippocampus.bl | $-1.970 \times 10^{-1}$ | $-3.970 \times 10^{-1}$ | $-7.330 \times 10^{-1}$ |
| FDG | $-7.470 \times 10^{-1}$ | $-4.030 \times 10^{-1}$ | -1.602 |
| FDG.bl | $-8.430 \times 10^{-1}$ | $-5.840 \times 10^{-1}$ | -1.487 |
| MOCA | $2.970 \times 10^{-1}$ | $-2.500 \times 10^{-2}$ | -1.302 |
| ABETA | 5.091 | 4.737 | -4.511 |
| PTAU | $-3.310 \times 10^{-1}$ | $4.620 \times 10^{-1}$ | 1.053 |
| TAU | $-8.590 \times 10^{-1}$ | 1.497 | 2.641 |
| AV45 | -1.464 | $-5.350 \times 10^{-1}$ | $-2.900 \times 10^{-1}$ |
| MOCA.bl | $-5.400 \times 10^{-1}$ | $-6.410 \times 10^{-1}$ | -1.343 |
| AV45.bl | -1.690 | $-7.460 \times 10^{-1}$ | $-5.370 \times 10^{-1}$ |
| imputed_genotype | -3.957 | -2.645 | -4.020 |
| ABETA.RAW | $1.838 \times 10^1$ | $1.727 \times 10^1$ | $-1.805 \times 10^1$ |
| PTAU.RAW | -8.429 | -1.502 | 1.270 |
| TAU.RAW | -8.692 | 2.586 | 6.368 |
| APOE Genotype | 0.000 | 0.000 | 0.000 |

**Table 9.**
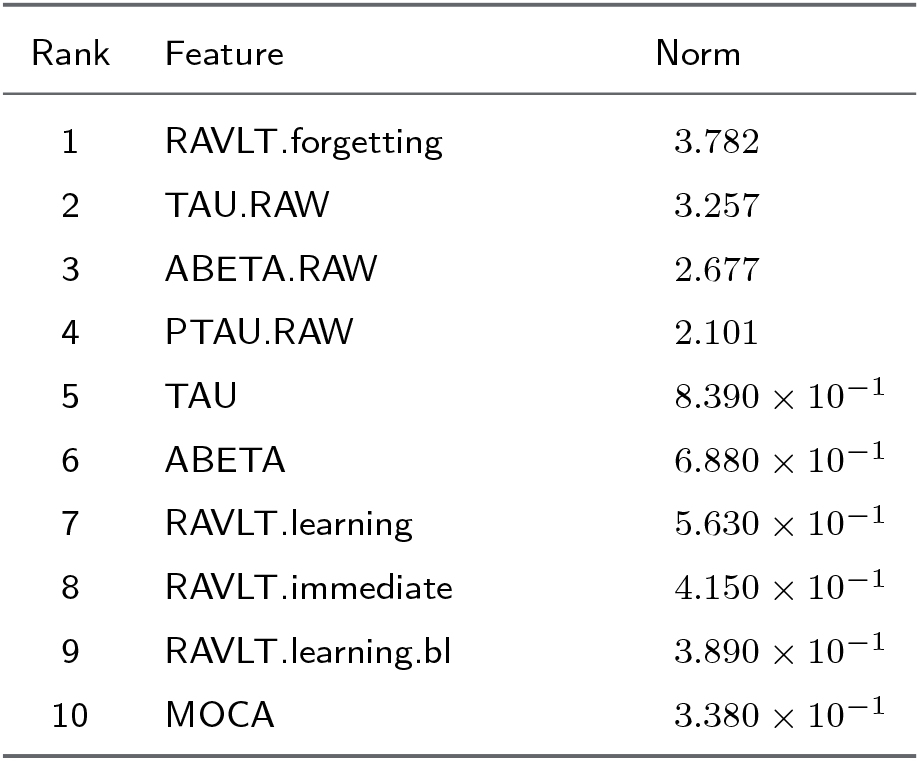
Top driver features ranked by norm values.

| Rank | Feature | Norm |
| --- | --- | --- |
| 1 | RAVLT.forgetting | 3.782 |
| 2 | TAU.RAW | 3.257 |
| 3 | ABETA.RAW | 2.677 |
| 4 | PTAU.RAW | 2.101 |
| 5 | TAU | $8.390 \times 10^{-1}$ |
| 6 | ABETA | $6.880 \times 10^{-1}$ |
| 7 | RAVLT.learning | $5.630 \times 10^{-1}$ |
| 8 | RAVLT.immediate | $4.150 \times 10^{-1}$ |
| 9 | RAVLT.learning.bl | $3.890 \times 10^{-1}$ |
| 10 | MOCA | $3.380 \times 10^{-1}$ |

**Table 10.** Feature-wise p-values from statistical tests across clusters.

| Feature | p-value |
| --- | --- |
| APOE A2 | $1.241 \times 10^{-22}$ |
| APOE A1 | $7.029 \times 10^{-12}$ |
| MMSE | $7.518 \times 10^{-31}$ |
| MMSE.bl | $6.822 \times 10^{-22}$ |
| RAVLT.forgetting | $1.464 \times 10^{-239}$ |
| RAVLT.learning | $1.333 \times 10^{-34}$ |
| RAVLT.immediate | $2.119 \times 10^{-44}$ |
| FAQ | $1.405 \times 10^{-42}$ |
| RAVLT.forgetting.bl | $1.244 \times 10^{-127}$ |
| RAVLT.learning.bl | $3.232 \times 10^{-28}$ |
| RAVLT.immediate.bl | $2.388 \times 10^{-37}$ |
| RAVLT.perc.forgetting | $1.122 \times 10^{-120}$ |
| FAQ.bl | $3.903 \times 10^{-32}$ |
| RAVLT.perc.forgetting.bl | $1.729 \times 10^{-85}$ |
| Hippocampus | $2.914 \times 10^{-15}$ |
| Hippocampus.bl | $1.087 \times 10^{-14}$ |
| FDG | $1.063 \times 10^{-22}$ |
| FDG.bl | $1.033 \times 10^{-23}$ |
| MOCA | $1.032 \times 10^{-37}$ |
| ABETA | $2.725 \times 10^{-64}$ |
| PTAU | $8.942 \times 10^{-50}$ |
| TAU | $2.385 \times 10^{-44}$ |
| AV45 | $6.972 \times 10^{-33}$ |
| MOCA.bl | $2.659 \times 10^{-32}$ |
| AV45.bl | $1.884 \times 10^{-32}$ |
| imputed_genotype | $9.978 \times 10^{-20}$ |
| ABETA.RAW | $1.187 \times 10^{-64}$ |
| PTAU.RAW | $1.525 \times 10^{-68}$ |
| TAU.RAW | $2.269 \times 10^{-59}$ |
| APOE Genotype | — |

